# Target engagement during interleaved iTBS-fMRI predicts subsequent clinical response to connectivity-guided accelerated iTBS: results of the PRISM-TRD trial

**DOI:** 10.64898/2026.04.09.26350062

**Authors:** Christopher B. Pople, Maria Vasileiadi, Alina Zaidi, Dara Silver, Lina Musa, Alexander J. Nyman, Anusha Baskaran, Enrique Collins-Rivera, Fa-Hsuan Lin, Robin F.H. Cash, Andrew Zalesky, Adriano Mollica, Maged Goubran, Katharine Dunlop, Robert Chen, Jamie Near, Muhammad Ishrat Husain, Jennifer S. Rabin, Daniel M. Blumberger, Benjamin Davidson, Clement Hamani, Peter Giacobbe, Nir Lipsman, Martin Tik, Sean M. Nestor

## Abstract

**Objective:** Reliable prognostic biomarkers in depression remain elusive. The objective of this study was to assess whether interleaved intermittent theta-burst stimulation-fMRI (iTBS-fMRI) can measure real-time target engagement, and prospectively test whether target engagement was associated with subsequent response to accelerated, connectivity-guided iTBS in treatment-resistant depression.

**Methods:** This single-centre trial (NCT05813093) included 71 patients (42 with treatment-resistant depression [TRD], 29 ultra-treatment-resistant [UTRD]) who received personalized left dorsolateral prefrontal cortex (dlPFC) targeting based on functional connectivity with the subgenual anterior cingulate cortex (sgACC). Participants underwent one sham-controlled iTBS-fMRI session followed by five days of open-label accelerated iTBS (40 sessions; 600 pulses each). The primary outcome was change in HAM-D_17_. Primary imaging analyses examined active-sham interleaved iTBS-fMRI effects in the sgACC and left dlPFC regions, and their relationship with clinical outcomes.

**Results:** Accelerated iTBS was associated with rapid, sustained symptom reductions (ΔHAM-D_17_ -9.01, p<0.001); overall response and remission rates were 40.8% (TRD: 47.6%, UTRD: 31.0%) and 16.9% (TRD: 16.7%, UTRD: 17.2%), with benefit maintained at 4 and 12 weeks. During interleaved iTBS-fMRI, active stimulation elicited significant target engagement within the sgACC-associated seedmap used for targeting (p=0.029). This brain response correlated with subsequent clinical improvement in depression and anxiety, but not other symptoms, and provided additional prognostic value independent of established clinical and demographic predictors.

**Conclusions:** Connectivity-guided accelerated iTBS was associated with rapid antidepressant effects in both TRD and UTRD participants with less pronounced effects in UTRD. Online target engagement measured by interleaved iTBS-fMRI is a candidate mechanistic biomarker that may aid prognosis in the future.

## Introduction

Major depressive disorder (MDD) is a debilitating illness affecting ∼20% of individuals during their lifetime (1). Among treatment-seeking patients, 30–50% do not respond to one or more antidepressant therapies and meet criteria for treatment-resistant depression (TRD) (2, 3). Greater chronicity and failure of interventional treatments, such as electroconvulsive therapy (ECT) may identify a more refractory subgroup (3), but ultra-treatment-resistant depression (UTRD) has no consensus definition.

TRD involves persistent mood, hedonic, and cognitive-affective disturbances alongside functional and structural alterations in frontocingulate networks (4–6). Repetitive transcranial magnetic stimulation (rTMS) noninvasively modulates these networks, inducing neuroplastic changes (5, 7). Intermittent theta burst stimulation (iTBS), an approved rTMS protocol, delivers high-frequency bursts at theta frequency (5 Hz), approximating plasticity induction protocols in preclinical models (8, 9). iTBS provides comparable benefits with shorter sessions than earlier protocols (3.5 vs. 20-40 minutes) (10). Nevertheless, response rates remain heterogeneous and are often below 50% (11).

The subgenual anterior cingulate cortex (sgACC) is a hub within mood-regulation networks that exhibits increased activity and connectivity in MDD (12–16), while the left dorsolateral prefrontal cortex (dlPFC) is often hypoactive in depression and has negative functional and effective connectivity with the sgACC (4, 14–19). Mechanistically diverse antidepressants are shown to reverse biological alterations of this frontocingulate circuit (6, 12–14, 17), and retrospective studies suggest that dlPFC-sgACC connectivity may predict antidepressant response to rTMS (14–16, 20). The US Food and Drug Administration recently cleared a protocol that implements connectivity-guided accelerated iTBS targeting personalized left dlPFC regions selected by sgACC anticorrelation (21).

Despite an improved neurobiological and circuit-based understanding of MDD, reliable prognostic biomarkers remain elusive (2, 22), potentially reflecting the correlational nature and limited mechanistic specificity of many candidate biomarkers, together with substantial interindividual variability in neurophysiological responses and treatment outcomes (23, 24). Interleaving TMS and fMRI (TMS-fMRI) combines the temporal resolution of TMS with the spatial resolution of fMRI to measure brain responses during perturbation across distributed networks (25, 26). Prior studies have associated acute brain responses during TMS-fMRI with subsequent clinical improvement (18, 19, 27) but lacked sham controls and used single-pulse TMS or 1 Hz right dlPFC rTMS delivered between fMRI volume acquisitions.

A standard 600-pulse iTBS pattern used in clinical care was recently delivered using interleaved TMS-fMRI (28), enabling measurement of perturbation responses to iTBS and other therapeutic rTMS protocols. This approach may support mechanistic studies and the development of prognostic biomarkers to predict therapeutic response to rTMS, guide rTMS protocol development, and clarify neurobiological mechanisms of TRD.

Here we prospectively delivered sham-controlled iTBS-fMRI at personalized, connectivity-guided targets before open-label connectivity-guided accelerated iTBS (cg-aiTBS) at the same targets in patients with TRD of varying refractoriness. This design let us assess online target engagement by iTBS in a clinical population and test its association with subsequent clinical improvement, as assessed by the HAM-D_17_, after repeated delivery of the same intervention. We expected that cg-aiTBS would produce rapid antidepressant effects and that activation of the dlPFC and deactivation of the sgACC would be associated with greater clinical response.

## Methods

### Study Design

This single-centre, open-label, pilot trial (NCT05813093), titled PRISM-TRD (Predicting Response via Interleaved intermittent theta-burst Stimulation-functional Magnetic resonance imaging (fMRI) in Treatment-Resistant Depression), investigated the relationship of pre-treatment neuroimaging biomarkers collected during interleaved rTMS-fMRI with subsequent clinical outcomes in a cohort of patients with TRD of heterogeneous refractoriness.

Participants completed baseline 3T structural MRI and resting-state fMRI (Figure 1). These data, combined with electric-field modeling, were used to derive an individualized left dlPFC stimulation target maximally anticorrelated with an sgACC-associated seedmap (see (20)). At a second visit, patients underwent neuronavigated TMS-fMRI (active and sham) at this target, before five days of open-label neuronavigated cg-aiTBS (8 sessions/day, 600 pulses/session) at the same target. Additional acquisition, clinical, and medication details are provided in the Supplemental Methods.

**Figure 1.**
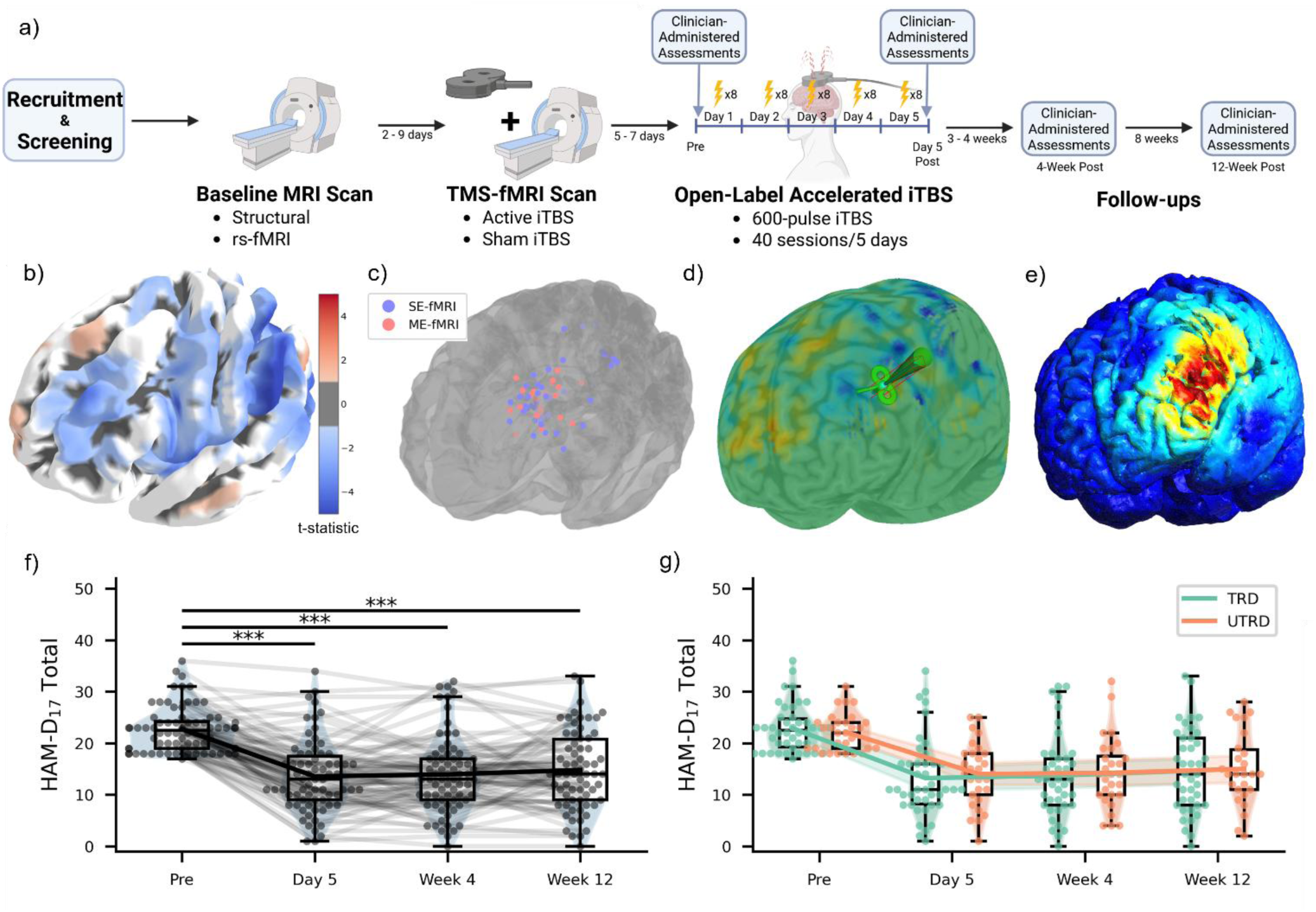
Study timeline, connectivity-guided targeting, and primary clinical outcomes. Top: a) Study timeline, Middle: b) group-mean resting-state functional connectivity (rsFC) surface map (darker reds represent strong positive rsFC, and darker blues represent stronger negative rsFC), c) Location of each subject’s rTMS targets derived from functional connectivity with an sgACC-associated seedmap displayed on the cortical surface (blue = single-echo fMRI, red = multi-echo fMRI), and d) a target coordinate in the dlPFC exhibiting greatest negative functional connectivity with the sgACC and a correlated seedmap visualized in Brainsight neuronavigation software (positive rsFC = yellow-red, negative rsFC = teal-blue), e) an E-field simulation (depicting magnitude in V/m; blue = zero, red = greatest intensity) was performed at that target to find an optimal coil trajectory, which was then imported into Brainsight for treatment delivery. Bottom: Change in HAM-D_17_ score, prior to rTMS treatment and again at follow-up visits. Panel (f) represents the entire cohort and panel (g) shows clinical response trajectories for TRD and UTRD groups. Lines track group mean, and box plots show median and interquartile ranges. Bonferroni correction was applied for repeated longitudinal comparisons before significance testing. * = p_adj_ < 0.05, ** = p_adj_ < 0.01, *** = p_adj_ < 0.001.

### Patient Recruitment

The Sunnybrook Health Sciences Centre Research Ethics Board approved the study, and all participants provided informed consent in accordance with the Declaration of Helsinki and institutional policies. Participants were recruited from the Harquail Centre for Neuromodulation between November 2023 through July 2025 through two clinical streams. The TRD group was recruited from an interventional psychiatry rTMS clinic, and the UTRD group (high refractoriness stream) was recruited from a psychiatric neurosurgery clinic evaluating candidates for investigational deep brain stimulation or focused ultrasound capsulotomy.

Participants were aged 20–65 and required a primary diagnosis of MDD or persistent depressive disorder, a current major depressive episode of at least moderate severity (grid-standardized HAM-D_17_ ≥18 measured by an interventional psychiatrist), and have not responded to ≥1 adequate antidepressant trial. Exclusion criteria included MRI contraindications, depression secondary to another primary psychiatric disorder, history of psychosis or seizure disorder, active pregnancy, and substance use disorder within the previous six months.

Participants were assigned *a priori* to protocol-defined subgroups: TRD or UTRD. The TRD subgroup required insufficient response to ≥1 adequate antidepressant trial (29). The UTRD subgroup required all the following: (i) illness duration ≥5 years; (ii–iii) >6 antidepressant trials and >1 adjunct trial of adequate dose and duration; (iv) ≥1 psychotherapy modality; and (v) prior ketamine, rTMS, or ECT. As UTRD is not a consensus category, Maudsley Staging Method (MSM) scores and individual treatment histories were reported to support interpretation.

### Baseline MRI and connectivity-guided targeting

The first 43 participants underwent single- and multi-echo resting-state fMRI (rs-fMRI) for a planned technical comparison. Based on an exploratory interim analysis (Supplemental Results) and considering prior evidence supporting multi-echo rs-fMRI (30), multi-echo data were used prospectively for target localization in the remaining 28 participants. Acquisition and sequence parameters are available in the Supplemental Methods. After acquisition-specific preprocessing and denoising, personalized rsFC targets were identified using a published pipeline (14), and SimNIBS-optimized coil trajectories were imported into Brainsight for neuronavigation. Acquisition, software, and pipeline details are provided in the Supplemental Methods.

### Interleaved TMS-fMRI acquisition

At the second MRI visit, interleaved TMS-fMRI data were acquired on a Siemens Prisma 3T system, using the posterior shell of a head/neck coil (20- or 64-channel), a 4-channel flexible body coil, and in later subjects, a 4-channel carotid imaging coil. We transitioned to the later coil configuration for greater tSNR (Suppl. Fig. 1), given the earlier configuration’s limited tSNR, particularly in the sgACC (Suppl. Fig. 2). Stimulation was delivered in the scanner using an MRi-B91 coil (MagVenture, Denmark) guided by optical neuronavigation, with trains of iTBS delivered between multi-band echo-planar slice acquisitions.

Sham-controlled iTBS-fMRI data were acquired with two MRI coil configurations throughout the study (Suppl. Fig. 3). A 1-cm plastic spacer (Suppl. Fig. 4) was used to reduce the marginally suprathreshold (100% rMT) intensity to a subthreshold, subclinical level (31, 32) while preserving sensory confounds such as scalp sensations (Suppl. Fig. 5). Full acquisition and sham procedures are described in the Supplemental Methods.

### Accelerated iTBS treatment protocol

Participants received 40 cg-aiTBS sessions over five days targeting the left dlPFC (8/day; 50-minute inter-session interval) titrated to 110% resting motor threshold. Treatment stimulation used a Cool-B70 TMS coil (MagVenture) positioned and maintained along the SimNIBS-optimized trajectory under neuronavigated guidance (Supplemental Methods).

### Clinical Outcome Measures

Clinician-administered assessments, the GRID-HAM-D_17_ (primary outcome) and Montgomery-Åsberg Depression Rating Scale (MADRS) were collected by an interventional psychiatrist at baseline, after day 5, and at 4 and 12 weeks following treatment. Response was defined as a ≥50% decline in score, and remission as GRID-HAM-D_17_ ≤7 and MADRS ≤9, based on thresholds validated against clinician-rated global illness severity (33, 34). Exploratory self-report scales were also collected at these visits, with a subset of scales collected daily during treatment (Supplemental Methods).

### Statistical Analysis

Clinical and demographic differences between TRD and UTRD subgroups were quantified by standardized mean difference (SMD). Primary clinician-administered outcome measures, as well as differences in outcomes between TRD and UTRD groups, were tested by t-test (paired and independent, respectively) without adjustment for multiple comparisons. Blinding during TMS-fMRI was assessed by binomial test of correct/incorrect guesses. First-level event-related brain responses during iTBS-fMRI were estimated voxelwise and averaged within a spherical sgACC ROI (16), a dlPFC E-field ROI, and the sgACC-associated seedmap used for targeting (20).

Mixed-effects models included a four-level fixed effect of time and subject-specific random intercepts and slopes for a binary post-treatment regressor. Planned contrasts compared baseline with the mean post-treatment outcome or covariate slope. Longitudinal contrasts were Bonferroni-adjusted across time, while contrasts of brain-response slopes across outcomes were controlled using the two-stage Benjamini-Hochberg procedure. Independent prognostic value of brain responses was assessed by analysis of deviance testing the fit of nested logistic regression models predicting response or remission using age, sex, baseline symptom severity, and treatment resistance (MSM) with or without brain responses as predictors. Significance was set at α < 0.05. Additional details of the statistical analysis are reported in the Supplemental Methods.

## Results

### Baseline demographics

Of 96 patients assessed for eligibility (Suppl. Fig. 6), 73 were enrolled, and 71 proceeded with treatment after two withdrew before the intervention (median age 41 years [range 27-65], 26M/45F; 42 TRD, 29 UTRD). All 71 completed a baseline MRI and underwent cg-aiTBS. Most received concomitant psychiatric medications and psychotherapy (Suppl. Table 1). Although this study initially intended to enroll 88 subjects, recruitment was halted before reaching this target due to funding limitations.

Groups were similar in age, sex, and clinician-rated depression severity. UTRD participants had greater anhedonia, self-reported suicidality, and illness duration. As expected, moderate-to-large differences in MSM score and prior pharmacological and neuromodulation treatments were present (Table 1).

**Table 1.** Baseline Characteristics. **Table 1.** Demographics and clinical characteristics of the full study cohort at baseline, as well as *a priori* treatment resistance-stratified subgroups, and differences between these groups (standardized mean difference). Motor threshold using the treatment rTMS coil (Cool B70) is shown. Continuous data normally distributed as assessed by the Shapiro-Wilk test are presented as mean (SD), and otherwise as median [IQR]. Response on the HAM-D_17_ and the MADRS reflected a ≥ 50% decrease in scores, where remission reflected a HAM-D_17_ ≤ 7 or a MADRS ≤ 9. Categorical variables are presented as percent (count/total). SD: standard deviation, IQR: interquartile range. ECT: electroconvulsive therapy. MSM: Maudsley Staging Method. SMD: standardized mean difference.

|  | Overall (N=71) | TRD (N=42) | UTRD (N=29) | SMD |
| --- | --- | --- | --- | --- |
| **Age (Years)** | 41.0 [33.5 - 54.5] | 45.0 [34.2 - 55.0] | 40.0 [33.0 - 52.0] | -0.043 |
| **Sex (% Male)** | 36.6% (26/71) | 40.5% (17/42) | 31.0% (9/29) | -0.198 |
| **Illness Duration (Years)** | 21.1 (12.1) | 20.0 (13.1) | 22.7 (10.5) | 0.225 |
| **Motor Threshold (% MSO)** | 41.0 [37.0 - 48.0] | 41.1 (7.5) | 43.8 (8.7) | 0.321 |
| **Treatment History** |  |  |  |  |
| Total (No.) | 8.0 [5.0 - 13.2] | 7.0 [4.0 - 9.0] | 14.0 [8.0 - 17.0] | 0.641 |
| Current Episode (No.) | 2.0 [1.0 - 4.0] | 2.0 [1.0 - 3.0] | 3.0 [1.0 - 7.0] | 0.286 |
| Psychotherapies (No.) | 4.0 [2.0 - 4.0] | 3.1 (1.7) | 3.6 (1.7) | 0.286 |
| Prior Ketamine (%) | 21.1% (15/71) | 4.8% (2/42) | 44.8% (13/29) | 1.047 |
| Prior rTMS (%) | 19.7% (14/71) | 2.4% (1/42) | 44.8% (13/29) | 1.154 |
| Prior ECT (%) | 12.7% (9/71) | 0.0% (0/42) | 31.0% (9/29) | 0.949 |
| MSM Score | 9.6 (1.9) | 9.0 [8.0 - 10.0] | 11.0 [9.0 - 12.0] | 0.521 |
| **HAM-D<sub>17</sub>** | 22.0 [19.0 - 24.0] | 22.5 [19.2 - 24.8] | 22.0 [19.0 - 24.0] | -0.042 |
| **MADRS** | 30.7 (5.4) | 31.0 (5.6) | 30.2 (5.1) | -0.151 |
| **BDI** | 30.7 (8.6) | 30.1 (9.6) | 31.6 (6.9) | 0.176 |
| **QIDS-SR-16** | 20.9 (5.2) | 20.6 (5.5) | 21.4 (4.7) | 0.157 |
| **PHQ-9** | 19.0 [14.5 - 22.0] | 18.0 [12.5 - 22.0] | 20.0 [17.0 - 21.0] | 0.135 |
| **BSS** | 8.0 [3.0 - 14.0] | 7.0 [1.0 - 12.0] | 11.0 [5.8 - 15.0] | 0.275 |
| **GAD-7** | 12.0 [8.0 - 15.5] | 11.9 (4.9) | 11.9 (4.9) | 0.086 |
| **BITe** | 15.0 [10.0 - 18.5] | 14.2 (4.3) | 14.7 (5.3) | 0.096 |
| **DARS** | 65.8 (19.9) | 69.4 (19.0) | 60.7 (20.3) | -0.444 |
| **TAS-20** | 55.3 (11.9) | 55.3 (11.8) | 55.2 (12.4) | -0.009 |

### Connectivity-guided targeting

At the group level, negative rsFC was observed throughout the anterior dlPFC (Fig. 1b), though individual rsFC maps often demonstrated local peaks of negative rsFC (Fig. 1d). Target locations were heterogeneous, tending to cluster around the anterior dlPFC (Fig. 1c; group mean MNI X = -39.5 mm, Y = 34.7 mm, Z = 29.3 mm). Among the first 43 participants, greater Euclidean distance between single- and multi-echo targets was associated with less improvement on the HAM-D_17_ (p=0.025) and MADRS (p=0.029). Multi-echo data were therefore used for target localization in the remaining 28 participants (Suppl. Fig. 7; Supplemental Results). Clinical outcomes did not differ significantly between targeting methods (p=0.403).

### Accelerated, connectivity-guided iTBS is associated with rapid and sustained clinical improvement regardless of treatment resistance

Connectivity-guided, accelerated iTBS was associated with rapid improvement in our primary outcome measure, the HAM-D_17_ (mean ΔHAM-D_17_ -9.01 points [SD 6.06], t = -12.45, p < 0.001) by the end of treatment, with comparable effects in TRD vs UTRD (mean ΔHAM-D_17_ - 9.64 points [SD 5.94] vs -8.10 [SD 6.12], t = -1.05, p = 0.299) (Table 2, Fig. 1f,g). Response and remission rates at the end of treatment were 40.8% and 16.9%, respectively, and were not significantly different between TRD (47.6% and 16.7%) and UTRD (31.0% and 17.2%). In sensitivity analyses, change in HAM-D_17_ at the end of treatment also did not significantly correlate with number of previous psychiatric medications (ρ = 0.136, p = 0.257) or MSM score (r = 0.015, p = 0.899). A contrast of marginal means comparing post-treatment timepoints vs baseline supported an overall reduction in HAM-D_17_ score (−8.42 [SE 0.68], d.f. = 69.93, t = - 12.32, p < 0.001) and individual post-hoc tests of day 5, week 4, and week 12 timepoints vs baseline were all significant (p_adj_ < 0.05). In a supporting analysis, MADRS scores showed similar sustained improvement without a TRD–UTRD group-wise difference (Table 2; Suppl. Fig. 8).

**Table 2.** Clinician-administered outcomes. **Table 2.** Clinician-administered outcome measures for the entire cohort as well as *a priori* treatment resistance-stratified subgroups and differences between these groups (statistic and p-value). Continuous data normally distributed as assessed by the Shapiro-Wilk test are presented as mean (SD) and differences tested by two-sample t-test. Otherwise, continuous variables are presented as median [IQR] and differences tested by Mann-Whitney U test. Categorical variables are presented as percent (count/total) and tested using Fisher’s exact test. SD: standard deviation, IQR: interquartile range. HAM-D_17_: Hamilton Depression Rating Scale, 17-item. MADRS: Montgomery-Åsberg Depression Rating Scale.

|  | Overall (N=71) | TRD (N=42) | UTRD (N=29) | statistic | p-value |
| --- | --- | --- | --- | --- | --- |
| **HAM-D<sub>17</sub>** |  |  |  |  |  |
| Pre, total | 22.0 [19.0 - 24.0] | 22.5 [19.2 - 24.8] | 22.0 [19.0 - 24.0] | 583.5 | 0.768 |
| Post, total | 13.6 (7.0) | 11.0 [8.2 - 16.0] | 14.0 [10.0 - 18.0] | 691.0 | 0.339 |
| Post, responders | 40.8% (29/71) | 47.6% (20/42) | 31.0% (9/29) | 0.495 | 0.221 |
| Post, remitted | 16.9% (12/71) | 16.7% (7/42) | 17.2% (5/29) | 1.042 | 1.000 |
| Week 4, total | 13.0 [9.0 - 17.0] | 13.0 [8.0 - 17.0] | 14.0 [10.0 - 17.5] | 619.5 | 0.582 |
| Week 4, responders | 37.7% (26/69) | 46.3% (19/41) | 25.0% (7/28) | 0.386 | 0.083 |
| Week 4, remitted | 17.4% (12/69) | 19.5% (8/41) | 14.3% (4/28) | 0.688 | 0.749 |
| Week 12, total | 14.8 (7.6) | 14.7 (8.1) | 15.0 (7.1) | 0.193 | 0.848 |
| Week 12, responders | 36.4% (24/66) | 40.0% (16/40) | 30.8% (8/26) | 0.667 | 0.601 |
| Week 12, remitted | 16.7% (11/66) | 20.0% (8/40) | 11.5% (3/26) | 0.522 | 0.505 |
| **MADRS** |  |  |  |  |  |
| Pre, total | 30.7 (5.4) | 31.0 (5.6) | 30.2 (5.1) | -0.629 | 0.531 |
| Post, total | 18.5 (9.8) | 17.7 (9.9) | 19.7 (9.6) | 0.86 | 0.393 |
| Post, responders | 39.4% (28/71) | 42.9% (18/42) | 34.5% (10/29) | 0.702 | 0.622 |
| Post, remitted | 15.5% (11/71) | 16.7% (7/42) | 13.8% (4/29) | 0.8 | 1.000 |
| Week 4, total | 18.0 [11.0 - 26.0] | 17.9 (10.8) | 20.1 (10.8) | 0.816 | 0.418 |
| Week 4, responders | 41.2% (28/68) | 50.0% (20/40) | 28.6% (8/28) | 0.4 | 0.087 |
| Week 4, remitted | 23.5% (16/68) | 25.0% (10/40) | 21.4% (6/28) | 0.818 | 0.780 |
| Week 12, total | 19.1 (10.0) | 18.7 (10.8) | 19.7 (8.9) | 0.433 | 0.667 |
| Week 12, responders | 33.3% (22/66) | 40.0% (16/40) | 23.1% (6/26) | 0.45 | 0.188 |
| Week 12, remitted | 22.7% (15/66) | 27.5% (11/40) | 15.4% (4/26) | 0.479 | 0.369 |

Self-reported depression, suicidality, and anxiety also significantly improved, whereas irritability improved less consistently and anhedonia and alexithymia did not change significantly (Suppl. Figs. 9 and 10; Suppl. Tables 2 and 3; Supplemental Results). In an exploratory analysis of the most densely sampled measure, the BDI, improvement was significant on the first day of treatment (mean ΔBDI = -1.65 points [SE 0.56], t = -2.94, d.f. = 68, p_adj_ = 0.044), and continued thereafter (Suppl. Fig. 9a; Suppl. Table 2). Change in self-reported outcomes did not differ between TRD and UTRD groups on any scale at any timepoint (Suppl. Fig. 10).

Treatment was well-tolerated with no withdrawals during open-label cg-aiTBS, and no severe or serious adverse events occurred. Adverse events were predominantly mild, transient, and temporally-associated with stimulation, most frequently scalp discomfort, headache, fatigue, and facial twitching (Suppl. Table 4; Suppl. Fig. 11; Supplemental Results). One patient withdrew prior to treatment due to MRI-related anxiety.

### iTBS-fMRI elicited target engagement in a distributed network of functionally-connected brain regions that was correlated with subsequent clinical improvement

The interleaved iTBS-fMRI cohort (Suppl. Fig. 6) comprised 57 patients with similar clinical and demographic characteristics (Suppl. Table 5), and clinician-rated outcomes (Suppl. Fig. 12, Suppl. Table 6). Interleaved iTBS-fMRI was generally well-tolerated, with mostly mild, exclusively transient adverse events. Self-reported pain and discomfort were collected for a subset of patients and were greater in the active than the sham condition (t = 5.96, d.f. = 37, p < 0.001), though 59.6% guessed the condition correctly (34/57, p = 0.185). Consistent with this, large motion spikes were common during the first train of iTBS (Suppl. Fig. 13; Supplemental Results), and were controlled with a 12-parameter motion model, ICA-AROMA non-aggressive denoising, and censoring of motion spikes and surrounding volumes (Supplemental Methods).

To assess target engagement, responses were tested both separately by condition and as an active-sham contrast. No significant event-related response in the *a priori* left dlPFC (thresholded E-field) and bilateral spherical sgACC ROIs (16) was detected in either condition, nor was an active-sham difference observed (Fig. 2a,b). As tSNR was low in these ROIs but higher throughout much of the rest of the brain (Suppl. Figs. 1 and 2), we also performed a post-hoc analysis of activation in the distributed sgACC-associated seedmap derived from the same sgACC ROI (20) and used *a priori* for targeting prior to interleaved iTBS-fMRI. Here, significant activation occurred in the active (t = 2.247, p = 0.029), but not sham condition (t = 1.270, p = 0.209) although the active-sham difference did not reach significance (t = 1.235, p = 0.222), potentially due to subthreshold effects in the sham condition (Fig. 2c).

**Figure 2.**
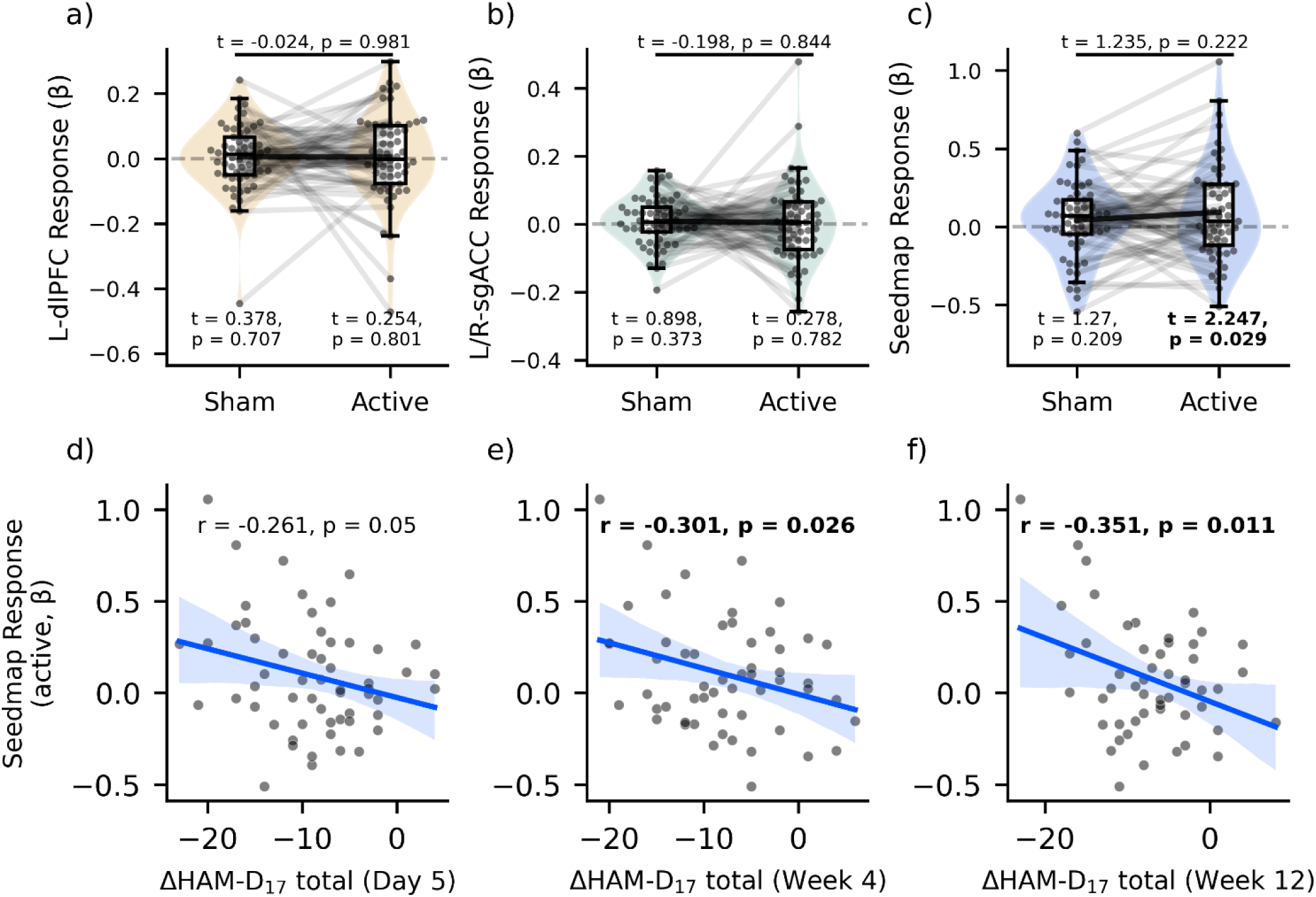
iTBS-fMRI elicits target engagement in an sgACC-associated network that is correlated with subsequent clinical improvement. Top row: Event-related response in a) the dlPFC E-field, b) Bilateral sgACC, and c) sgACC-associated seedmap, in active and sham conditions. Bottom row: Pearson’s correlation of event-related responses in sgACC-associated seedmap in the active condition with subsequent change in HAM-D_17_ at day 5, week 4, and week 12.

We next tested whether seedmap engagement predicted subsequent clinical outcomes. Mixed-effects modelling revealed a significant standardized association (β_Time X ROI_ = -0.231 [SE 0.096], d.f. = 55.15, t = -2.41, p = 0.019) between active condition seedmap responses and improvement in HAM-D_17_ after cg-aiTBS. In sensitivity analyses, this relationship remained significant after adjusting for age, sex, and MSM score (p = 0.019), as well as when adjusting for targeting methodology (single-vs multi-echo rs-fMRI) and coil configuration (p = 0.019). Univariate correlations similarly showed a trend at day 5 with significant correlation at weeks 4 and 12 (Fig. 2d-f). Conversely, this relationship was much weaker in the sham condition, only reaching significance for week 12, and was absent in the active-sham contrast (Suppl. Fig. 14). As in activation analyses, no relationship was observed between change in HAM-D_17_ and sgACC or left dlPFC responses in either condition or the contrast (Suppl. Fig. 15).

### Seedmap engagement during iTBS-fMRI provides within-sample prognostic value independent of clinical and demographic predictors that generalizes across outcome measures

Next, we examined whether the prognostic value of iTBS-fMRI seedmap responses extended beyond the HAM-D_17_. Active-condition iTBS-fMRI seedmap responses were significantly associated (Fig. 3a) with subsequent improvement on the HAM-D_17_, PHQ-9, and QIDS-SR-16 depression scales, as well as anxiety (GAD-7), and a trend for MADRS. However, no relationship was observed with suicidality (BSS), irritability (BITe), anhedonia (DARS) or alexithymia (TAS-20). In sensitivity analyses adjusting for i) age, sex and MSM, as well as ii) targeting methodology and coil configuration, results were comparable except the relationship with MADRS survived FDR correction (data not shown). Although a similar pattern of non-significant correlations was apparent in seedmap responses in the sham condition, no significant associations were observed in other ROIs and conditions.

**Figure 3.**
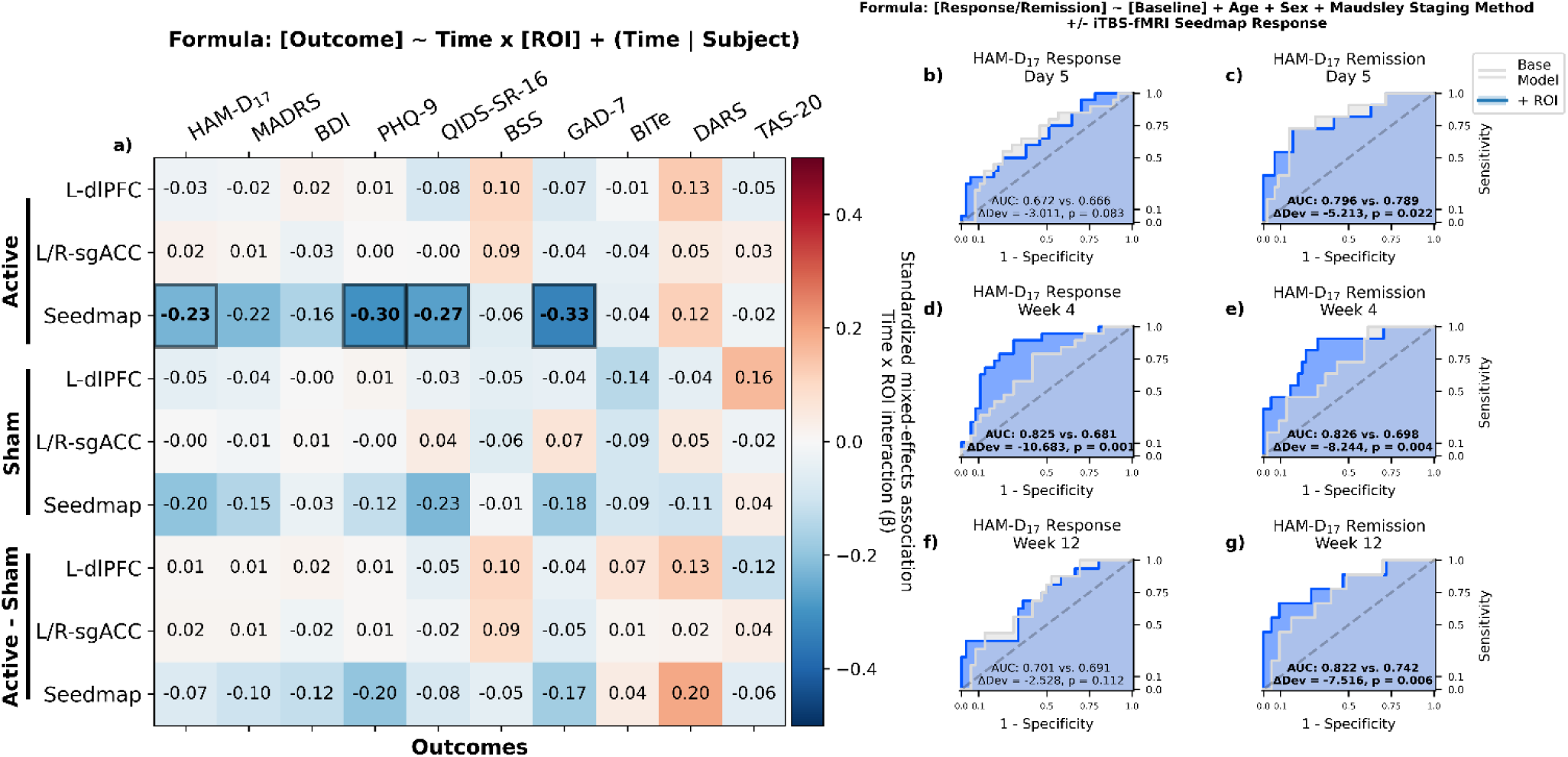
Robust prognostic value of brain responses during iTBS-fMRI is orthogonal to clinical and demographic predictors. Left: a) Standardized mixed-effects associations between iTBS-fMRI responses and longitudinal change in clinician-administered and self-reported psychiatric scales were modeled using a four-level categorical fixed effect of time (baseline, day 5, week 4, and week 12), with subject-specific random intercepts and slopes for a binary post-treatment regressor. Bolded: p_FDR_ < 0.05 after two-stage Benjamini-Hochberg adjustment for the number of symptom scores tested (i.e. within-row). Right: (b-g) Receiver-operator curves demonstrating predictive capacity of nested models, testing whether logistic regression models including (blue) event-related responses during iTBS-fMRI have independent prognostic value relative to a base model (white) including demographic (age, sex) and clinical variables (baseline symptom severity, MSM). Prediction of response (b,d,f) and remission (c,e,g) are shown separately at day 5 (b,c), week 4 (d,e), and week 12 (f,g) on the HAM-D_17_. Bolded: Analysis of deviance p < 0.05.

To test whether iTBS-fMRI responses added prognostic information above and beyond clinical and demographic variables, we compared logistic regression models predicting HAM-D_17_ response and remission from age, sex, baseline symptom severity, and MSM score, with or without seedmap responses in the active condition (Fig. 3b-g). Nested model comparison by analysis of deviance assessed whether brain responses added incremental within-sample prognostic value beyond clinical and demographic variables previously associated with rTMS outcomes (35–37). Within-sample HAM-D_17_ remission prediction was significantly improved at every timepoint (Fig. 3c,e,g), as was response prediction at week 4 (Fig. 3d), but not day 5 and week 12 (Fig. 3b,f). MADRS response and remission prediction was uniformly improved by seedmap responses at every timepoint (Suppl. Fig. 16). Sham condition responses offered weaker, less consistent prognostic value (Suppl. Fig. 17). Consistent with univariate correlations, responses in the bilateral sgACC (Suppl. Fig. 18) and left dlPFC (Suppl. Fig. 19) added no apparent within-sample prognostic value.

## Discussion

We prospectively tested whether online target engagement measured with interleaved 600-pulse iTBS-fMRI was associated with clinical outcomes after sequential accelerated cg-aiTBS neuronavigated to the same target in TRD. To our knowledge, this was the largest single-cohort TMS-fMRI study to date. Treatment at personalized left dlPFC targets defined by connectivity with an sgACC-associated seed map (20) was associated with rapid, sustained improvement in both TRD and UTRD groups, although reductions in depression scores were numerically smaller in UTRD. Active, but not sham, iTBS-fMRI significantly engaged the distributed targeting seed map, although the active–sham contrast was nonsignificant. Pre-treatment active, but not sham, iTBS-fMRI engagement of the distributed targeting seed map was associated with subsequent reductions in depression and anxiety scale scores with cg-aiTBS, and active iTBS-fMRI added within-sample prognostic value above clinical and demographic variables previously associated with rTMS outcomes (35–37). These findings also generalized to self-reported depression and anxiety scale scores but no other symptom domains.

Personalized rsFC-based targets were broadly distributed across the left dlPFC but concentrated anterolaterally, consistent with previously reported effective targets (14–16). Multi-echo targets occupied a somewhat smaller area than single-echo fMRI sequence-derived targets, potentially reflecting more consistent rsFC estimation (30, 38), but clinical outcomes were similar.

Although response and remission rates were lower than those initially reported for Stanford Neuromodulation Therapy (SNT) (sgACC connectivity, 1800-pulse iTBS, 10 sessions/day), protocol, sample, and outcome-definition differences limit direct comparison (21). By 12 weeks, reported response and remission rates after SNT had declined to levels comparable to those observed here (39). In both cases rapid antidepressant effects emerged over the 5-day acute course. This suggests that cg-aiTBS may be appropriate in TRD even with long illness duration and many prior treatment failures, although this study was not powered to test non-inferiority in UTRD.

The sham iTBS-fMRI control closely resembled active stimulation and helped to disambiguate iTBS/rTMS-induced effects from auditory and somatosensory stimuli that confounded interpretation in previous TMS-fMRI studies in depression (18, 19, 27). E-field simulations suggested our sham condition was equivalent to ≤80% rMT stimulation, a level previously reported to be clinically ineffective (31, 32); however, it may have produced subthreshold activity and altered excitability (40–42). Indeed, seed map activation and clinical relationships in the sham condition were of lower magnitude but similar direction and pattern as those observed in the active condition, and effects were stronger than those observed in other tested ROIs and conditions.

Contrary to our *a priori* hypothesis, we did not observe significant dlPFC or sgACC ROI responses during active and sham iTBS-fMRI, nor ROI relationships with clinical outcomes. Several previous TMS-fMRI studies similarly failed to detect activation at the stimulation site (18, 19, 43). In contrast, a healthy control study detected dlPFC responses at a lower rMT (80%) (28), while studies using similar coil configurations reported inconsistent local dlPFC activation (44, 45). The absence of apparent sgACC engagement is likely attributable to poor sgACC tSNR, inherent with imaging deeper brain structures with concurrent TMS-fMRI versus conventional fMRI setups. For both ROIs, our power was also limited by the brief acquisition during iTBS, high motion often time-locked with stimuli, and the long TR (2 s) of our fMRI sequence.

TMS-fMRI is an emerging method for causal mapping of brain network function and connectivity, as well as a source of prognostic biomarkers in psychiatry and beyond (25, 26). Our results are consistent with previous studies that demonstrated relationships between brain responses to active, single-pulse TMS and clinical response (18, 19, 27). We extend these findings with one of the largest interleaved TMS-fMRI studies to date, utilizing a sham-controlled design and a therapeutic iTBS protocol in a cohort that included individuals with highly treatment-refractory illnesses. Furthermore, we show that target engagement during iTBS-fMRI, in the same sgACC-associated seedmap used for connectivity-guided targeting, is associated with subsequent clinical improvement to accelerated iTBS.

## Limitations & Future Directions

Our findings should be interpreted with the following limitations. Open-label treatment precluded separating iTBS-specific from nonspecific or placebo-related improvement, and heterogeneous concomitant medications may have influenced fMRI and clinical responses. Low tSNR inherent with TMS-fMRI setups, subthreshold sham effects, brief acquisition, and stimulus-associated motion further limited sensitivity and active-sham differences. Multi-echo targeting and our second MRI coil configuration were introduced nonrandomly during enrollment following an exploratory interim analysis and consideration of prior technical evidence. Consequently, the study was not designed to determine whether multi-echo targeting improves clinical efficacy, and comparisons between targeting methods may be affected by temporal, procedural, or other cohort differences. Lastly, while we demonstrated within-sample prognostic value of iTBS-fMRI responses, generalizability is unclear given the lack of a replication cohort or pre-specified thresholds. Future studies should improve acquisition sensitivity through optimized coil configurations, longer stimulation/scanning protocols, better motion control, and fMRI sequences offering more frequent sampling and/or higher tSNR.

## Supporting information

Supplemental Results

## Data Availability

All data produced in the present study are available upon reasonable request to the authors, pursuant to legal and institutional regulations and guidelines.

## Acknowledgements

We acknowledge our MRI technologists, Ruby Endre and Sankyu Moon, for their substantial support in neuroimaging data collection as well as with interventions and procedures conducted during these visits.

## Notes

### Competing Interest Statement

This work was funded by Sean Nestor's 2022 NARSAD Young Investigator Grant from the Brain & Behavior Research Foundation. Additional funding support was provided by the Labatt Family Discovery Program of the Labatt Family Network for Research on the Biology of Depression, Department of Psychiatry, Temerty Faculty of Medicine, University of Toronto, and the Harquail Centre for Neuromodulation, Sunnybrook Research Institute, and Sunnybrook Research Foundation provided funding for the rTMS equipment.
Fa-Hsuan Lin receives research support from Natural Sciences and Engineering Research Council of Canada (NSERC; RGPIN-2020-05927), Canada Foundation for Innovation (38913 and 41351), Canadian Institutes of Health Research (PJT 178345 and PJT 496433), and MITACS (IT25405).
Daniel M Blumberger receives research support from CIHR, NIMH, Wellcome Trust, Brain Canada and the Temerty Family through the CAMH Foundation and the Campbell Family Research Institute, has received research grant support and in-kind equipment support for an investigator-initiated study from Brainsway Ltd, was the site principal investigator for three sponsor-initiated studies for Brainsway Ltd, has received in-kind equipment support from Magventure for investigator-initiated studies, has received medication supplies for an investigator-initiated trial from Indivior, is a scientific advisor for Sooma Medical, is a member of the Clinical Standards Committee of the Clinical TMS Society (unpaid) and is the Provincial Clinical Lead for neurostimulation procedures for Ontario Health (paid).
Clement Hamani is a consultant for Abbott, and his research is supported by a Tier 1 Canada Research Chair in Translational Research.
Andrew Zalesky is supported by the ARC Future Fellowship and Rebecca L. Cooper Foundation, and is an executive member of the Queensland Neurostimulation Centre.
Robin F.H. Cash is supported by the Australian National Health and Medical Research Council (Emerging Leadership Investigator grant no. 2017527).
Robert Chen receives research support from CIHR, NSERC, Parkinson Foundation, Weston Brain Institute and was a consultant for Abbvie, Merz, Ipsen and NovusTx.
Katharine Dunlop receives salary support from an Academic Scholars Award, Department of Psychiatry, University of Toronto. Katharine Dunlop also receives research support from CIHR, Brain Canada, the American Foundation for Suicide Prevention Early Career Research Award, Labatt Family Network Catalyst Grant, and the Brain & Behaviour Research Foundation Young Investigator Grant.

### Clinical Trial

NCT05813093

### Author Declarations

This study was approved by Sunnybrook Health Sciences Centre's Research Ethics Board (protocol ID 5408) on July 7, 2022.

### Summary of Updates

Revision ahead of journal submission to align with submission format/constraints, moving some results to a supplement, reduced citation count, and contextual editing to reduce word count while preserving content. Integrated some feedback and corrected some inconsistencies in methods/captions that emerged between revisions. Minor corrections to author affiliations.

### Funding

- Brain & Behavior Research Foundation: 2022 NARSAD Young Investigator Grant
- Labatt Family Network for Research on the Biology of Depression: Labatt Family Discovery Program
- Department of Psychiatry, Temerty Faculty of Medicine, University of Toronto
- Harquail Centre for Neuromodulation, Sunnybrook Research Institute, and Sunnybrook Research Foundation

## References

1. Hasin DS, Sarvet AL, Meyers JL, et al.: Epidemiology of Adult DSM-5 Major Depressive Disorder and Its Specifiers in the United States. JAMA Psychiatry 2018; 75:336–346

2. Warden D, Rush AJ, Trivedi MH, et al.: The STAR*D Project results: a comprehensive review of findings. Curr Psychiatry Rep 2007; 9:449–459

3. McIntyre RS, Alsuwaidan M, Baune BT, et al.: Treatment-resistant depression: definition, prevalence, detection, management, and investigational interventions. World Psychiatry 2023; 22:394–412

4. Pizzagalli DA, Roberts AC: Prefrontal cortex and depression. Neuropsychopharmacol 2022; 47:225–246

5. Nestor SM, Mir-Moghtadaei A, Vila-Rodriguez F, et al.: Large-scale structural network change correlates with clinical response to rTMS in depression. Neuropsychopharmacol 2022; 47:1096– 1105

6. Siddiqi SH, Schaper FLWVJ, Horn A, et al.: Brain stimulation and brain lesions converge on common causal circuits in neuropsychiatric disease. Nat Hum Behav 2021; 5:1707–1716

7. Lee C-W, Wu H-F, Chu M-C, et al.: Mechanism of Intermittent Theta-Burst Stimulation in Synaptic Pathology in the Prefrontal Cortex in an Antidepressant-Resistant Depression Rat Model. Cerebral Cortex 2021; 31:575–590

8. Larson J, Munkácsy E: Theta-burst LTP. Brain Research 2015; 1621:38–50

9. Nuñez A, Buño W: The Theta Rhythm of the Hippocampus: From Neuronal and Circuit Mechanisms to Behavior. Front Cell Neurosci 2021; 15:649262

10. Blumberger DM, Vila-Rodriguez F, Thorpe KE, et al.: Effectiveness of theta burst versus high-frequency repetitive transcranial magnetic stimulation in patients with depression (THREE-D): a randomised non-inferiority trial. The Lancet 2018; 391:1683–1692

11. Vida RG, Sághy E, Bella R, et al.: Efficacy of repetitive transcranial magnetic stimulation (rTMS) adjunctive therapy for major depressive disorder (MDD) after two antidepressant treatment failures: meta-analysis of randomized sham-controlled trials. BMC Psychiatry 2023; 23:545

12. Alexander L, Jelen LA, Mehta MA, et al.: The anterior cingulate cortex as a key locus of ketamine’s antidepressant action. Neuroscience & Biobehavioral Reviews 2021; 127:531–554

13. Hamani C, Mayberg H, Stone S, et al.: The Subcallosal Cingulate Gyrus in the Context of Major Depression. Biological Psychiatry 2011; 69:301–308

14. Cash RFH, Cocchi L, Lv J, et al.: Functional Magnetic Resonance Imaging–Guided Personalization of Transcranial Magnetic Stimulation Treatment for Depression. JAMA Psychiatry 2021; 78:337– 339

15. Weigand A, Horn A, Caballero R, et al.: Prospective Validation That Subgenual Connectivity Predicts Antidepressant Efficacy of Transcranial Magnetic Stimulation Sites. Biological Psychiatry 2018; 84:28–37

16. Fox MD, Buckner RL, White MP, et al.: Efficacy of Transcranial Magnetic Stimulation Targets for Depression Is Related to Intrinsic Functional Connectivity with the Subgenual Cingulate. Biological Psychiatry 2012; 72:595–603

17. Fales CL, Barch DM, Rundle MM, et al.: Antidepressant treatment normalizes hypoactivity in dorsolateral prefrontal cortex during emotional interference processing in major depression. Journal of Affective Disorders 2009; 112:206–211

18. Oathes DJ, Zimmerman JP, Duprat R, et al.: Resting fMRI-guided TMS results in subcortical and brain network modulation indexed by interleaved TMS/fMRI. Exp Brain Res 2021; 239:1165–1178

19. Oathes DJ, Duprat RJ-P, Reber J, et al.: Non-invasively targeting, probing and modulating a deep brain circuit for depression alleviation. Nat Mental Health 2023; 1:1033–1042

20. Cash RFH, Cocchi L, Lv J, et al.: Personalized connectivity-guided DLPFC-TMS for depression: Advancing computational feasibility, precision and reproducibility. Human Brain Mapping 2021; 42:4155–4172

21. Cole EJ, Phillips AL, Bentzley BS, et al.: Stanford Neuromodulation Therapy (SNT): A Double-Blind Randomized Controlled Trial. AJP 2022; 179:132–141

22. Abi-Dargham A, Moeller SJ, Ali F, et al.: Candidate biomarkers in psychiatric disorders: state of the field. World Psychiatry 2023; 22:236–262

23. Corp DT, Bereznicki HGK, Clark GM, et al.: Large-scale analysis of interindividual variability in theta-burst stimulation data: Results from the ‘Big TMS Data Collaboration.’ Brain Stimulation 2020; 13:1476–1488

24. Hinchman CA, Fried PJ, Jannati A, et al.: Corticomotor plasticity as a predictor of response to high frequency transcranial magnetic stimulation treatment for major depressive disorder. Journal of Affective Disorders 2022; 303:114–122

25. Siddiqi SH, Kording KP, Parvizi J, et al.: Causal mapping of human brain function. Nat Rev Neurosci 2022; 23:361–375

26. Bergmann TO, Varatheeswaran R, Hanlon CA, et al.: Concurrent TMS-fMRI for causal network perturbation and proof of target engagement. NeuroImage 2021; 237:118093

27. Ge R, Humaira A, Gregory E, et al.: Predictive Value of Acute Neuroplastic Response to rTMS in Treatment Outcome in Depression: A Concurrent TMS-fMRI Trial. AJP 2022; 179:500–508

28. Chang K-Y, Tik M, Mizutani-Tiebel Y, et al.: Neural response during prefrontal theta burst stimulation: Interleaved TMS-fMRI of full iTBS protocols. NeuroImage 2024; 291:120596

29. Milev RV, Giacobbe P, Kennedy SH, et al.: Canadian Network for Mood and Anxiety Treatments (CANMAT) 2016 Clinical Guidelines for the Management of Adults with Major Depressive Disorder: Section 4. Neurostimulation Treatments. Can J Psychiatry 2016; 61:561–575

30. Lynch CJ, Elbau I, Liston C: Improving precision functional mapping routines with multi-echo fMRI. Current Opinion in Behavioral Sciences 2021; 40:113–119

31. Boutros NN, Gueorguieva R, Hoffman RE, et al.: Lack of a therapeutic effect of a 2-week sub-threshold transcranial magnetic stimulation course for treatment-resistant depression. Psychiatry Research 2002; 113:245–254

32. Padberg F, Zwanzger P, Keck ME, et al.: Repetitive Transcranial Magnetic Stimulation (rTMS) in Major Depression: Relation between Efficacy and Stimulation Intensity. Neuropsychopharmacol 2002; 27:638–645

33. Hawley CJ, Gale TM, Sivakumaran T: Defining remission by cut off score on the MADRS: selecting the optimal value. Journal of Affective Disorders 2002; 72:177–184

34. Leucht S, Fennema H, Engel R, et al.: What does the HAMD mean? J Affect Disord 2013; 148:243– 248

35. Lisanby SH, Husain MM, Rosenquist PB, et al.: Daily Left Prefrontal Repetitive Transcranial Magnetic Stimulation in the Acute Treatment of Major Depression: Clinical Predictors of Outcome in a Multisite, Randomized Controlled Clinical Trial. Neuropsychopharmacol 2009; 34:522–534

36. Trevizol AP, Downar J, Vila-Rodriguez F, et al.: Predictors of remission after repetitive transcranial magnetic stimulation for the treatment of major depressive disorder: An analysis from the randomised non-inferiority THREE-D trial. EClinicalMedicine 2020; 22:100349

37. Slan AR, Citrenbaum C, Corlier J, et al.: The role of sex and age in the differential efficacy of 10 Hz and intermittent Theta-burst (iTBS) repetitive transcranial magnetic stimulation (rTMS) treatment of major depressive disorder (MDD) [Internet]. Journal of Affective Disorders 2024; Available from: https://www.sciencedirect.com/science/article/pii/S0165032724013582

38. Hassanzadeh E, Tariq R, Palm S, et al.: Improved Reliability of Resting-State Functional MRI Connectivity Using Multi-Echo Acquisition: Implications for Personalized Transcranial Magnetic Stimulation Targeting [Internet]. American Journal of Neuroradiology 2026; [cited 2026 Mar 24] Available from: https://www.ajnr.org/content/early/2026/03/19/ajnr.A9301

39. Geoly AD, Stimpson KH, Espil FM, et al.: Durability of clinical benefit with Stanford Neuromodulation Therapy (SNT) in treatment-resistant depression. Brain Stimulation 2025; 18:875–881

40. Bestmann S, Baudewig J, Siebner HR, et al.: Subthreshold high-frequency TMS of human primary motor cortex modulates interconnected frontal motor areas as detected by interleaved fMRI-TMS. NeuroImage 2003; 20:1685–1696

41. Rollnik JD, Schubert M, Dengler R: Subthreshold prefrontal repetitive transcranial magnetic stimulation reduces motor cortex excitability. Muscle & Nerve 2000; 23:112–114

42. Tang AD, Bennett W, Bindoff AD, et al.: Subthreshold repetitive transcranial magnetic stimulation drives structural synaptic plasticity in the young and aged motor cortex. Brain Stimulation 2021; 14:1498–1507

43. Rafiei F, Rahnev D: TMS Does Not Increase BOLD Activity at the Site of Stimulation: A Review of All Concurrent TMS-fMRI Studies [Internet]. eNeuro 2022; 9[cited 2024 Mar 22] Available from: https://www.eneuro.org/content/9/4/ENEURO.0163-22.2022

44. Goldstein S, Rafiei F, Rahnev D: 3D-printed stand, timing interface, and coil localization tools for concurrent TMS-fMRI experiments. Brain Stimulation 2022; 15:1290–1291

45. Woolgar A, Feredoes E, Assem M, et al.: Consensus guidelines for the use of concurrent TMS-fMRI in cognitive and clinical neuroscience. Nat Protoc 2025; 1–17

