## Supplemental Materials for "Target engagement during interleaved iTBS-fMRI predicts subsequent clinical response to connectivity-guided accelerated iTBS: results of the PRISM-TRD trial"

**Supplemental Table 1. Concomitant psychiatric medications**

| Medication/Drug Class | Patients Receiving, N (%) |
| --- | --- |
| SSRIs | 29 / 71 (40.8%) |
| SNRIs | 14 (19.7%) |
| Tricyclic Antidepressants | 1 (1.4%) |
| Bupropion | 15 (21.1%) |
| Mirtazapine | 9 (12.7%) |
| Trazodone | 9 (12.7%) |
| Stimulants | 18 (26.8%) |
| Mood Stabilizers | 5 (7.0%) |
| Antipsychotics | 17 (23.9%) |
| Benzos & Z-Drugs | 20 (28.2%) |
| Gabapentinoids | 3 (4.2%) |
| MAOIs | 1 (1.4%) |

**Supplemental Table 1.** Concomitant psychiatric medications taken by patients during all interventions, and up to at least 4-weeks of follow-up. Numbers reflect unique patients taking the indicated drug or a drug from the indicated class, and may not add up to the total cohort due to many patients taking multiple medications. SSRIs: selective serotonin reuptake inhibitors. SNRIs: serotonin-norepinephrine reuptake inhibitors. MAOIs: monoamine oxidase inhibitors.

### Supplemental Table 2. Self-reported outcome measures

| | **Overall (N=71)** | **TRD (N=42)** | **UTRD (N=29)** | statistic | p-value |
|---|---|---|---|---|---|
| **BDI** | | | | | |
| Pre, total | 30.7 (8.6) | 30.1 (9.6) | 31.6 (6.9) | 0.752 | 0.455 |
| Day 1, total | 28.7 (9.4) | 28.2 (10.0) | 29.3 (8.6) | 0.499 | 0.620 |
| Day 2, total | 26.6 (10.8) | 26.4 (11.5) | 26.9 (10.0) | 0.192 | 0.848 |
| Day 3, total | 25.5 (11.6) | 24.6 (12.7) | 26.8 (9.9) | 0.798 | 0.427 |
| Day 4, total | 23.0 [16.0 - 33.0] | 21.0 [15.0 - 33.0] | 25.0 [18.0 - 33.0] | 650.5 | 0.631 |
| Post, total | 23.8 (12.5) | 20.0 [14.0 - 32.0] | 22.0 [17.5 - 34.0] | 590.0 | 0.411 |
| Week 1, total | 22.0 [15.0 - 34.0] | 23.1 (13.0) | 24.2 (10.3) | 0.357 | 0.723 |
| Week 2, total | 22.7 (12.1) | 20.5 (13.4) | 25.6 (9.8) | 1.699 | 0.095 |
| Week 3, total | 23.1 (11.2) | 21.0 (12.2) | 25.8 (9.4) | 1.701 | 0.094 |
| Week 4, total | 23.7 (11.7) | 22.2 (12.4) | 25.8 (10.4) | 1.243 | 0.219 |
| Week 12, total | 24.3 (12.0) | 22.5 (12.8) | 26.7 (10.6) | 1.392 | 0.169 |
| **PHQ-9** | | | | | |
| Pre, total | 19.0 [14.5 - 22.0] | 18.0 [12.5 - 22.0] | 20.0 [17.0 - 21.0] | 691.5 | 0.336 |
| Post, total | 13.6 (6.8) | 12.7 (6.8) | 15.0 (6.8) | 1.358 | 0.180 |
| Week 4, total | 13.2 (6.8) | 11.7 (6.5) | 15.4 (6.8) | 2.139 | **0.037** |
| Week 12, total | 13.4 (7.5) | 11.4 (7.4) | 15.9 (6.9) | 2.457 | **0.017** |
| **QIDS-SR-16** | | | | | |
| Pre, total | 20.9 (5.2) | 20.6 (5.5) | 21.4 (4.7) | 0.658 | 0.513 |
| Post, total | 14.5 [10.0 - 21.0] | 14.5 (6.5) | 17.1 (6.7) | 1.55 | 0.127 |
| Week 4, total | 14.6 (6.4) | 13.8 (6.3) | 15.8 (6.6) | 1.166 | 0.249 |
| Week 12, total | 14.5 (6.2) | 13.2 (6.0) | 16.1 (6.3) | 1.869 | 0.067 |
| **BSS** | | | | | |
| Pre, total | 8.0 [3.0 - 14.0] | 7.0 [1.0 - 12.0] | 11.0 [5.8 - 15.0] | 732.0 | 0.054 |
| Post, total | 3.0 [1.0 - 11.2] | 2.0 [0.2 - 9.2] | 4.5 [1.0 - 14.2] | 603.0 | 0.135 |
| Week 4, total | 3.0 [1.0 - 11.0] | 2.0 [0.0 - 8.0] | 5.0 [3.0 - 14.0] | 605.5 | **0.022** |
| Week 12, total | 4.0 [1.0 - 11.8] | 2.0 [0.0 - 8.0] | 7.0 [1.0 - 12.0] | 581.5 | 0.121 |
| **GAD-7** | | | | | |
| Pre, total | 12.0 [8.0 - 15.5] | 11.7 (4.9) | 12.1 (5.0) | 0.355 | 0.724 |
| Post, total | 8.7 (5.0) | 8.8 (5.2) | 8.5 (4.8) | -0.25 | 0.803 |
| Week 4, total | 9.0 [4.0 - 13.0] | 7.0 [4.0 - 12.0] | 9.0 [5.0 - 13.0] | 489.5 | 0.566 |
| Week 12, total | 9.2 (5.5) | 8.8 (5.5) | 9.8 (5.5) | 0.716 | 0.477 |
| **BITe** | | | | | |
| Pre, total | 15.0 [10.0 - 18.5] | 14.2 (4.3) | 14.7 (5.3) | 0.392 | 0.696 |
| Post, total | 11.0 [9.0 - 17.0] | 11.0 [9.0 - 15.0] | 11.0 [9.0 - 17.0] | 526.0 | 1.000 |
| Week 4, total | 10.0 [9.0 - 17.0] | 10.0 [8.2 - 14.8] | 12.0 [9.0 - 18.0] | 491.5 | 0.309 |
| Week 12, total | 12.0 [9.0 - 17.8] | 12.9 (4.8) | 13.0 (4.9) | 0.146 | 0.885 |
| **DARS** | | | | | |
| Pre, total | 65.8 (19.9) | 69.4 (19.0) | 60.7 (20.3) | -1.829 | 0.073 |
| Post, total | 64.0 [56.0 - 85.0] | 73.7 (21.0) | 68.6 (24.6) | -0.863 | 0.392 |
| Week 4, total | 71.1 (23.6) | 72.9 (22.5) | 68.6 (25.3) | -0.668 | 0.507 |
| Week 12, total | 63.0 [51.0 - 87.0] | 64.0 [53.0 - 96.8] | 63.0 [50.0 - 80.0] | 413.0 | 0.509 |
| **TAS-20** | | | | | |
| Pre, total | 55.3 (11.9) | 55.3 (11.8) | 55.2 (12.4) | -0.035 | 0.972 |
| Post, total | 50.0 [45.0 - 61.8] | 52.9 (12.5) | 53.9 (11.8) | 0.339 | 0.736 |
| Week 4, total | 54.6 (11.1) | 54.4 (10.6) | 54.9 (12.0) | 0.165 | 0.870 |
| Week 12, total | 54.0 (12.3) | 53.7 (12.8) | 54.4 (11.8) | 0.221 | 0.826 |

**Supplemental Table 2.** Self-reported outcome measures for the entire cohort as well as *a priori* treatment resistance-stratified subgroups and differences between these groups (statistic and p-value). Continuous data normally distributed as assessed by the Shapiro-Wilk's test is presented as mean (SD) and differences tested by two-sample t-test. Otherwise, continuous variables are presented as median [IQR] and differences tested by Mann-Whitney U test. Categorical variables are presented as percent (count/total) and tested using Fisher's exact test. SD: standard deviation, IQR: interquartile range. BDI: Beck Depression Inventory. QIDS-SR-16: Quick Inventory of Depressive Symptomatology, Short Form, 16-item. PHQ-9: Patient Health Questionnaire, 9-item. BSS: Beck Scale for Suicidal Ideation. GAD-7: Generalized Anxiety Disorder scale, 7-item. BITe: Brief Irritability Test. DARS: Dimensional Anhedonia Rating Scale. TAS-20: Toronto Alexithymia Scale, 20-item.

**Supplemental Table 3. Overall effect of treatment contrast from mixed-effects models of longitudinal change in clinical scales.**

| Scale | Estimate | S.E. | d.f. | t-statistic | P-value (adjusted) |
|---|---|---|---|---|---|
| **Depression** | | | | | |
| *HAM-D$_{17}$* | -8.42 | 0.68 | 69.93 | -12.32 | **3.369e-18** |
| *MADRS* | -11.83 | 1.01 | 69.94 | -11.70 | **3.895e-17** |
| *BDI* | -6.66 | 0.92 | 66.85 | -7.20 | **6.643e-09** |
| *PHQ-9* | -4.63 | 0.66 | 67.27 | -7.01 | **1.455e-08** |
| *QIDS-SR-16* | -6.00 | 0.69 | 67.93 | -8.65 | **1.490e-11** |
| **Suicidality** | | | | | |
| *BSS* | -2.60 | 0.52 | 65.50 | -4.98 | **5.000e-05** |
| **Anxiety** | | | | | |
| *GAD-7* | -2.90 | 0.52 | 68.00 | -5.43 | **8.163e-06** |
| **Irritability** | | | | | |
| *BITe* | -1.44 | 0.42 | 67.75 | -3.38 | **0.012** |
| **Anhedonia** | | | | | |
| *DARS, total* | 3.83 | 2.04 | 67.34 | 1.88 | 0.651 |
| **Alexithymia** | | | | | |
| *TAS-20, total* | -1.85 | 0.82 | 67.62 | -2.26 | 0.270 |

**Supplemental Table 3.** Contrast estimates testing a post-treatment (day 5, week 4, week 12) vs pre-treatment estimated marginal means derived from a linear mixed effects models fitting a multi-level categorical effect of time to clinical scales. This reflects an overall treatment effect across follow-up timepoints. Scales are organized by symptoms, with models predicting scores on that scale longitudinally. From left to right, columns for each scale represent regressor name, point estimate (β), standard error (S.E.) of the estimate, degrees of freedom (d.f.), t-statistic, and Bonferroni-adjusted p-value. Degrees of freedom were calculated by the Kenward-Roger method. All models included a binary categorical random effect of time (i.e. Post-TMS) and intercept at the subject-level, consistent with subsequent models testing cross-level interactions. P-values significant at a threshold of $\alpha = 0.05$ are bolded. HAM-D$_{17}$: Hamilton Depression Rating Scale, 17-item. MADRS: Montgomery-Asberg Depression Rating Scale. BDI: Beck Depression Inventory. QIDS-SR-16: Quick Inventory of Depressive Symptomatology, Short Form, 16-item. PHQ-9: Patient Health Questionnaire, 9-item. BSS: Beck Scale for Suicidal Ideation. GAD-7: Generalized Anxiety Disorder scale, 7-item. BITe: Brief Irritability Test. DARS: Dimensional Anhedonia Rating Scale. TAS-20: Toronto Alexithymia Scale, 20-item.

### Supplemental Table 4. Summary of adverse events by relation to intervention

| Adverse Event | MRi-B91 Coil | | | Cool B70 A/P Coil | | | Unrelated | | |
|---|---|---|---|---|---|---|---|---|---|
| | No. Mild | No. Moderate | No. Patients | No. Mild | No. Moderate | No. Patients | No. Mild | No. Moderate | No. Patients |
| Facial twitching | 18 | 0 | 18 | 27 | 2 | 14 | 0 | 0 | 0 |
| Headache | 9 | 0 | 9 | 42 | 6 | 25 | 2 | 0 | 2 |
| Jaw discomfort | 0 | 0 | 0 | 6 | 3 | 6 | 0 | 0 | 0 |
| Lacrimation | 0 | 0 | 0 | 4 | 0 | 2 | 0 | 0 | 0 |
| Nausea | 7 | 7 | 0 | 13 | 1 | 10 | 1 | 1 | 2 |
| Scalp discomfort | 28 | 11 | 37 | 141 | 14 | 47 | 1 | 0 | 1 |
| Dizziness | 1 | 0 | 1 | 3 | 1 | 4 | 0 | 0 | 0 |
| Paraesthesia | 0 | 0 | 0 | 1 | 0 | 1 | 0 | 0 | 0 |
| Muscle twitching | 0 | 0 | 0 | 8 | 0 | 2 | 2 | 0 | 2 |
| Fatigue | 0 | 0 | 0 | 28 | 5 | 18 | 6 | 0 | 5 |
| Visual aura | 1 | 0 | 1 | 1 | 0 | 1 | 0 | 0 | 0 |
| Insomnia | 0 | 0 | 0 | 6 | 0 | 6 | 0 | 0 | 0 |
| Disorientation | 0 | 0 | 0 | 1 | 0 | 1 | 0 | 0 | 0 |
| Vomiting | 0 | 0 | 0 | 1 | 0 | 1 | 2 | 2 | 1 |
| Brain fog | 0 | 0 | 0 | 0 | 0 | 0 | 1 | 0 | 1 |
| Blurred vision | 0 | 0 | 0 | 0 | 0 | 0 | 1 | 0 | 1 |
| Gastrointestinal illness | 0 | 0 | 0 | 0 | 0 | 0 | 0 | 1 | 1 |
| Elevated hbA1c | 0 | 0 | 0 | 0 | 0 | 0 | 2 | 0 | 2 |
| Paresthesia | 0 | 0 | 0 | 0 | 0 | 0 | 1 | 0 | 1 |
| Intrusive thoughts | 0 | 0 | 0 | 0 | 0 | 0 | 0 | 1 | 1 |
| Anxiety | 0 | 0 | 0 | 0 | 0 | 0 | 5 | 2 | 4 |
| Hypertension | 0 | 0 | 0 | 0 | 0 | 0 | 1 | 0 | 1 |
| Dissociation | 0 | 0 | 0 | 0 | 0 | 0 | 0 | 1 | 1 |
| Light sensitivity | 0 | 0 | 0 | 0 | 0 | 0 | 1 | 0 | 1 |
| Influenza | 0 | 0 | 0 | 0 | 0 | 0 | 1 | 0 | 1 |
| Chest pain | 0 | 0 | 0 | 0 | 0 | 0 | 1 | 0 | 1 |
| Diarrhea | 0 | 0 | 0 | 0 | 0 | 0 | 1 | 0 | 1 |

**Supplemental Table 4.** Adverse Events reported between study entry and exit, organized by relationship with intervention and the Cool-B70 A/P rTMS coil used during the week of accelerated treatment. No severe adverse events were observed.

### Supplemental Table 5. Baseline Characteristics of the TMS-fMRI cohort

| | **Overall (N=57)** | **TRD (N=34)** | **UTRD (N=23)** | SMD |
|---|---|---|---|---|
| **Age (Years)** | 45.0 [34.0 - 55.0] | 46.0 [35.0 - 55.8] | 40.0 [33.0 - 53.0] | -0.064 |
| **Sex (% Male)** | 40.4% (23/57) | 41.2% (14/34) | 39.1% (9/23) | -0.042 |
| **Illness Duration (Years)** | 20.7 (11.6) | 19.7 (12.6) | 22.4 (9.7) | 0.238 |
| **Motor Threshold (% MSO)** | | | | |
| MRi-B91 | 62.6 (9.8) | 61.3 (9.4) | 64.6 (10.3) | 0.336 |
| Cool B70 | 41.0 [37.0 - 48.0] | 41.3 (7.2) | 43.7 (8.7) | 0.292 |
| **Treatment History** | | | | |
| Total (No.) | 8.0 [6.0 - 14.0] | 7.0 [4.2 - 9.0] | 14.0 [8.5 - 16.5] | 0.627 |
| Current Episode (No.) | 2.0 [1.0 - 4.0] | 2.0 [1.0 - 3.8] | 3.0 [1.0 - 7.0] | 0.145 |
| Psychotherapies (No.) | 4.0 [2.0 - 4.0] | 3.4 (1.6) | 3.4 (1.8) | 0.023 |
| Prior Ketamine (%) | 21.1% (12/57) | 5.9% (2/34) | 43.5% (11/23) | 0.969 |
| Prior rTMS (%) | 21.1% (12/57) | 2.9% (1/34) | 47.8% (11/23) | 1.204 |
| Prior ECT (%) | 10.5% (6/57) | 0.0% (0/34) | 26.1% (6/23) | 0.840 |
| MSM Score | 9.6 (1.8) | 9.0 [8.0 - 10.0] | 11.0 [9.0 - 12.0] | 0.451 |
| **HAM-D$_{17}$** | 22.0 [19.0 - 24.0] | 22.0 [18.2 - 26.5] | 22.0 [19.0 - 24.0] | -0.054 |
| **MADRS** | 30.9 (5.6) | 31.1 (6.0) | 30.5 (4.9) | -0.114 |
| **BDI** | 30.9 (8.7) | 30.5 (9.6) | 31.4 (7.4) | 0.112 |
| **QIDS-SR-16** | 20.8 (5.6) | 20.2 (5.9) | 21.6 (5.1) | 0.247 |
| **PHQ-9** | 17.9 (5.4) | 17.1 (6.0) | 18.9 (4.1) | 0.343 |
| **BSS** | 7.0 [2.0 - 14.0] | 6.5 [1.0 - 12.5] | 11.0 [6.0 – 16.0] | 0.307 |
| **GAD-7** | 12.0 [8.0 - 15.0] | 11.6 (4.6) | 12.2 (5.4) | 0.122 |
| **BITe** | 14.4 (4.6) | 14.5 (4.3) | 14.2 (5.1) | -0.063 |
| **DARS** | 66.2 (20.3) | 71.3 (19.4) | 58.7 (19.6) | -0.649 |
| **TAS-20** | 55.0 (11.8) | 53.9 (11.1) | 56.6 (12.8) | 0.230 |

**Supplemental Table 5.** Demographics and clinical characteristics of the iTBS-fMRI cohort at baseline, as well as *a priori* treatment resistance-stratified subgroups, and differences between these groups (standardized mean difference). Motor threshold using the treatment rTMS coil (Cool B70) is shown. Continuous data normally distributed as assessed by the Shapiro-Wilk test is presented as mean (SD), and otherwise as median [IQR]. Response on the HAM-D$_{17}$ and the MADRS reflected a ≥ 50% decrease in scores, where remission reflected a HAM-D$_{17}$ ≤ 7 or a MADRS ≤ 9. Categorical variables are presented as percent (count/total). SD: standard deviation, IQR: interquartile range. ECT: electroconvulsive therapy. MSM: Maudsley Staging Method. SMD: standardized mean difference

### Supplemental Table 6. Clinician-administered outcomes in the TMS-fMRI cohort

| | **Overall (N=57)** | **TRD (N=34)** | **UTRD (N=23)** | SMD |
|---|---|---|---|---|
| **$HAM\text{-}D_{17}$** | | | | |
| Pre, total | 22.0 [19.0 - 24.0] | 22.0 [18.2 - 26.5] | 22.0 [19.0 - 24.0] | -0.054 |
| Post, total | 13.6 (7.2) | 11.0 [8.2 - 16.0] | 14.0 [10.5 - 18.5] | 0.150 |
| Post, responders | 35.1% (20/57) | 41.2% (14/34) | 26.1% (6/23) | -0.324 |
| Post, remitted | 19.3% (11/57) | 20.6% (7/34) | 17.4% (4/23) | -0.082 |
| Week 4, total | 13.0 [9.0 - 18.5] | 14.4 (8.6) | 14.4 (7.7) | -0.007 |
| Week 4, responders | 34.5% (19/55) | 39.4% (13/33) | 27.3% (6/22) | -0.259 |
| Week 4, remitted | 20.0% (11/55) | 21.2% (7/33) | 18.2% (4/22) | -0.076 |
| Week 12, total | 15.1 (8.0) | 15.4 (8.6) | 14.8 (7.2) | -0.070 |
| Week 12, responders | 30.8% (16/52) | 34.4% (11/32) | 25.0% (5/20) | -0.206 |
| Week 12, remitted | 17.3% (9/52) | 21.9% (7/32) | 10.0% (2/20) | -0.329 |
| **MADRS** | | | | |
| Pre, total | 30.9 (5.6) | 31.1 (6.0) | 30.5 (4.9) | -0.114 |
| Post, total | 18.8 (10.1) | 17.9 (10.2) | 20.3 (9.9) | 0.236 |
| Post, responders | 36.8% (21/57) | 38.2% (13/34) | 34.8% (8/23) | -0.072 |
| Post, remitted | 17.5% (10/57) | 20.6% (7/34) | 13.0% (3/23) | -0.203 |
| Week 4, total | 19.0 [10.0 - 27.0] | 18.9 (11.4) | 20.3 (11.8) | 0.117 |
| Week 4, responders | 38.2% (21/55) | 42.4% (14/33) | 31.8% (7/22) | -0.221 |
| Week 4, remitted | 25.5% (14/55) | 24.2% (8/33) | 27.3% (6/22) | 0.069 |
| Week 12, total | 19.4 (10.3) | 19.2 (11.5) | 19.8 (8.5) | 0.066 |
| Week 12, responders | 26.9% (14/52) | 34.4% (11/32) | 15.0% (3/20) | -0.461 |
| Week 12, remitted | 21.2% (11/52) | 28.1% (9/32) | 10.0% (2/20) | -0.474 |

**Supplemental Table 6.** Clinician-administered outcome measures for the TMS-fMRI cohort as well as *a priori* treatment resistance-stratified subgroups and differences between these groups (statistic and p-value). Continuous data normally distributed as assessed by the Shapiro-Wilk's test is presented as mean (SD) and differences tested by two-sample t-test. Otherwise, continuous variables are presented as median [IQR] and differences tested by Mann-Whitney U test. Categorical variables are presented as percent (count/total) and tested using Fisher's exact test. SD: standard deviation, IQR: interquartile range. $HAM\text{-}D_{17}$: Hamilton Depression Rating Scale, 17-item. MADRS: Montgomery-Asberg Depression Rating Scale.

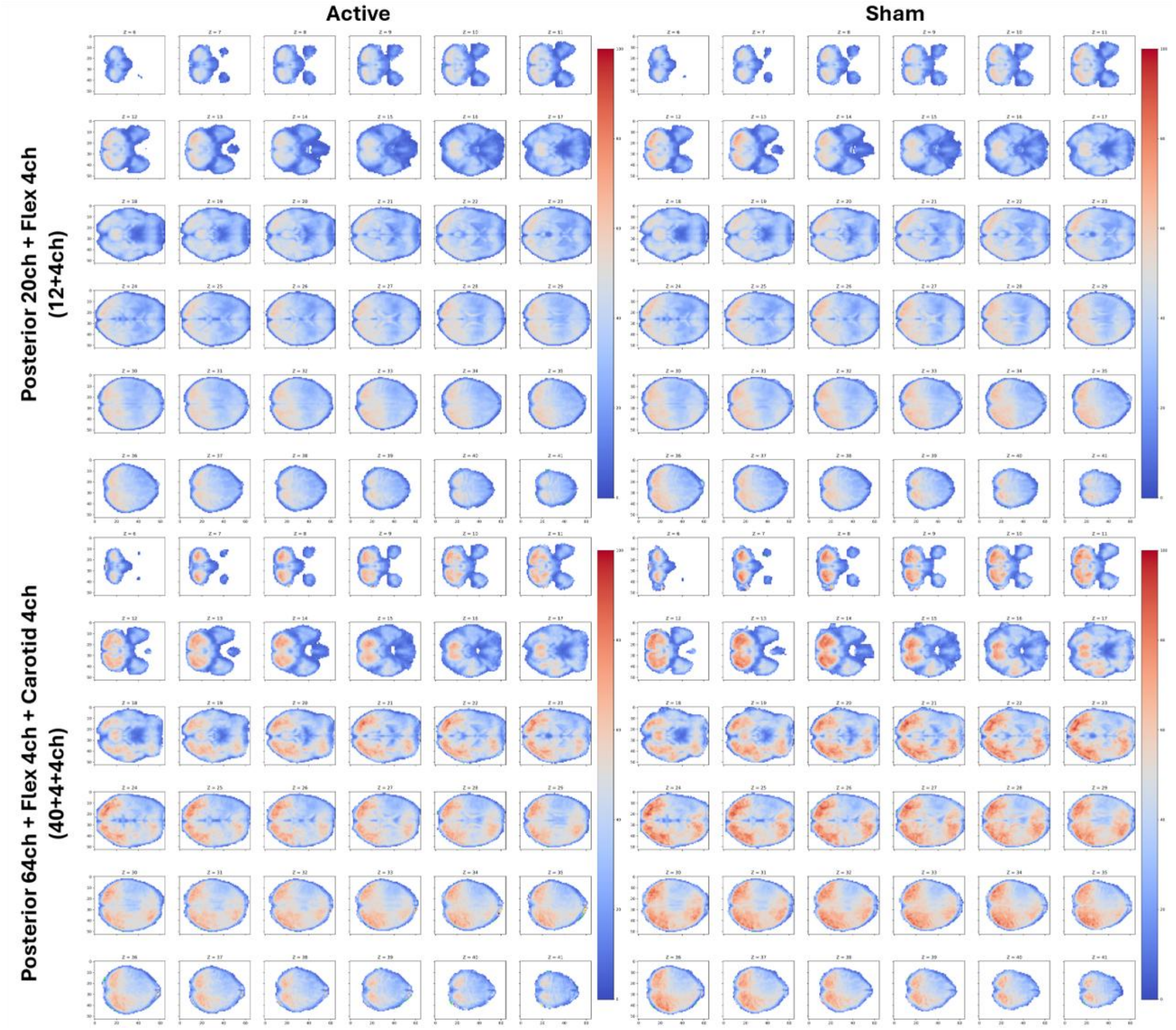


**Supplemental Figure 1. Temporal signal-to-noise ratio by condition and coil configuration.** Temporal signal-to-noise ratio maps in the (left) active and (right) condition for all subjects who had TMS-fMRI scans with the posterior shell of the 20-channel Siemens head coil and 4-channel flex body coil (top), and those who were scanned with the posterior shell of a 64-channel head coil, 4-channel flex coil, and 4-channel carotid coil on the opposite/right side of the head (against the rTMS coil).

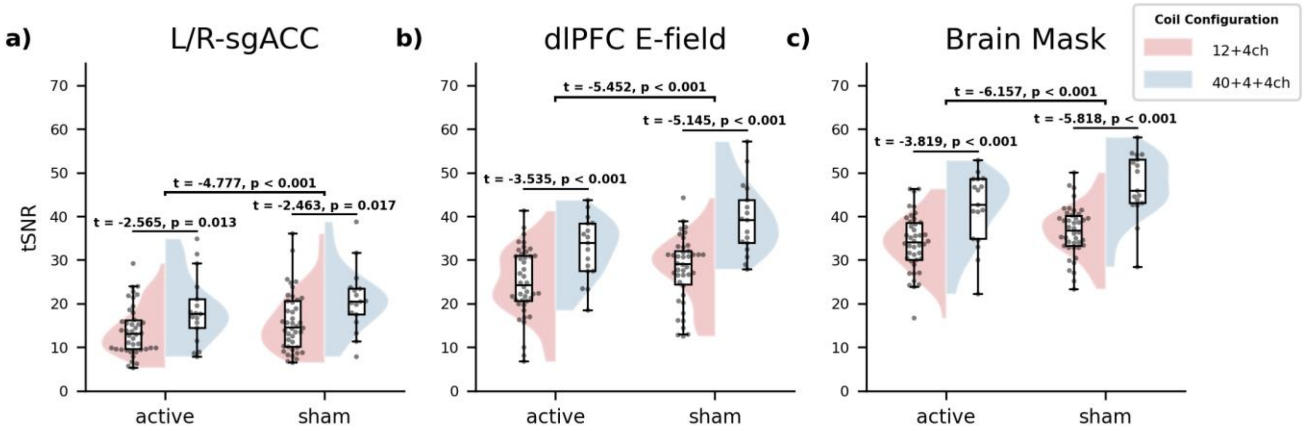


**Supplemental Figure 2. Mean temporal signal-to-noise ratio in each subjects ROI, by condition and coil configuration.** Temporal signal-to-noise ratio (tSNR) within a) a bilateral spherical sgACC ROI, b) each subject's dlPFC E-field, and c) the entire brain. In each case, tSNR is significantly higher in the sham condition, possibly due to lesser discomfort and less motion, and also significantly greater with the coil configuration with more channels. Notably, tSNR within the sgACC ROI was particularly.

**Contact corresponding author for figure**

**Supplemental Figure 3. TMS-fMRI data acquisition methodology.** The B91 MR compatible TMS coil (i) is positioned on the participant's head while lying on a vacuum head cushion (Siemens, Germany; 04765462), (ii) placed in the posterior shell of a 20-channel MR head/neck coil (Siemens, Germany) (iii) using a MagVenture TMS coil holder (iv) with six degrees of freedom. Optical trackers are placed on the subject (v) and TMS coil (vi) to allow optical neuronavigation (Rogue Research, Canada) with a Vega optical camera (Northern Digital, Canada) (vii). After the TMS coil is positioned and secured by the holder, the vacuum head fixation device (ii) is secured, and a flexible 4-channel MR radiofrequency body coil is placed over the TMS coil and the subject's head (inset). Two MRI coil configurations were used throughout this study. For the final 16 participants in the TMS-fMRI cohort, the posterior shell of a 64-channel head coil was used instead and ix) a 4-channel carotid coil (Siemens, Germany) was placed against the right side of the head opposite the rTMS coil prior to securing the 4-channel flex body coil. The participant is then transitioned into the scanner bore. During sham iTBS-fMRI procedure a 1 cm plastic spacer was covertly placed between the coil and the participant's head, outside of their field of view, immediately before locking the TMS coil into position.

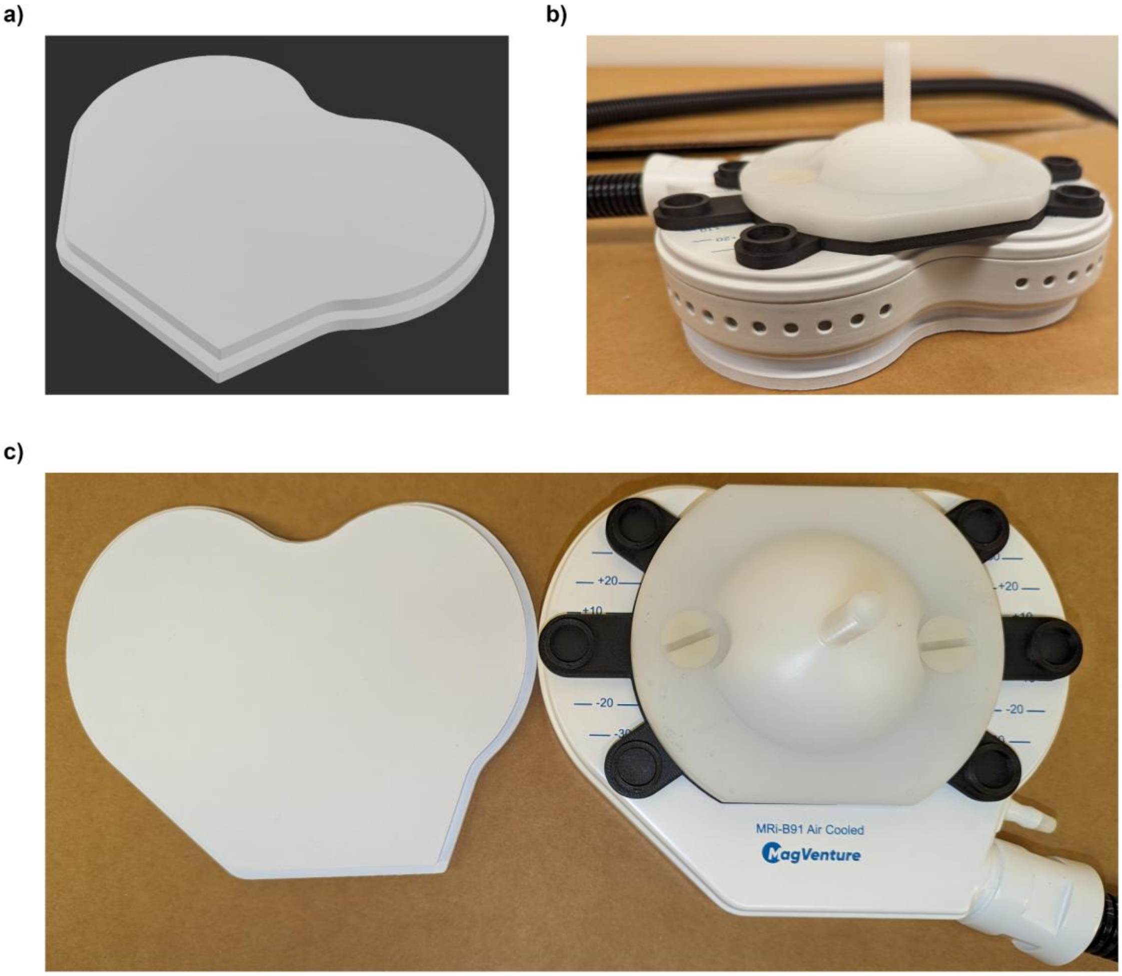


**Supplemental Figure 4. 3D-printed insert for sham-controlled TMS-fMRI.** 3D printed plastic spacer used to create sham condition. Render of 3D model (a), which was printed in polylactic acid (PLA) to create a 1cm thick insert designed to fit underneath the MRi-B91 TMS coil without protruding (b) owing to a similar but slightly smaller silhouette (c). The plastic spacer was slipped between the patient's head, secured firmly against the coil with surgical tape to prevent slipping, and the coil holder was locked with the spacer pressed against the head and coil.

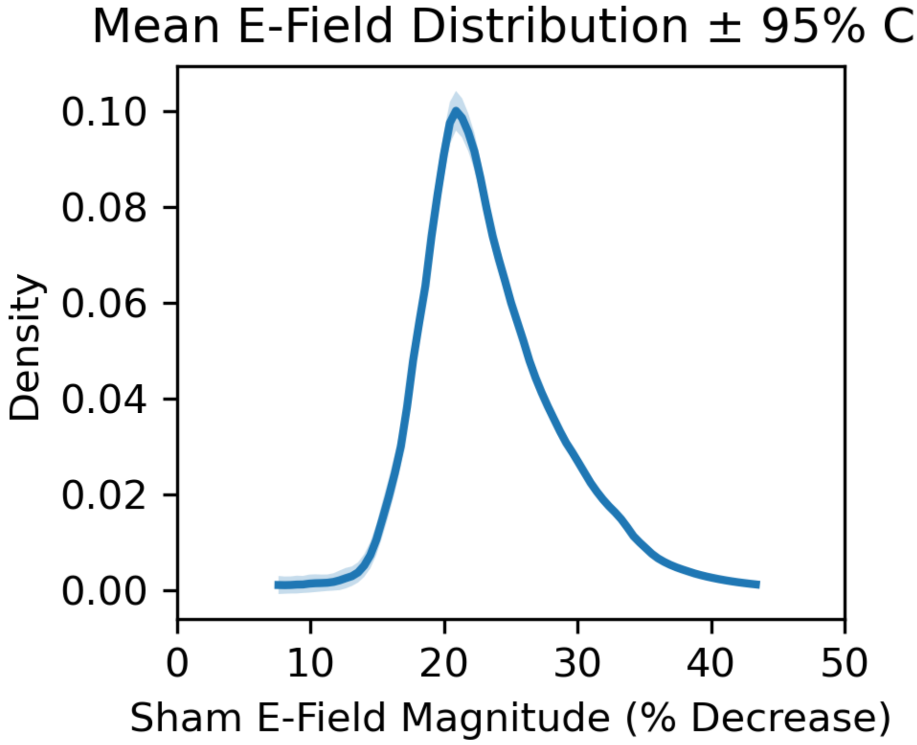


**Supplemental Figure 5. Relative decrease of E-Field magnitude during sham iTBS.** Mean histogram depicting distribution of (predicted) relative voxelwise E-field magnitude in the sham condition (1cm spacer inserted between coil and scalp) in each subject's dlPFC E-field ROI as predicted by the ratio of E-field magnitude when coil origin is displaced by 1cm from the scalp (sham) vs directly against it (active).

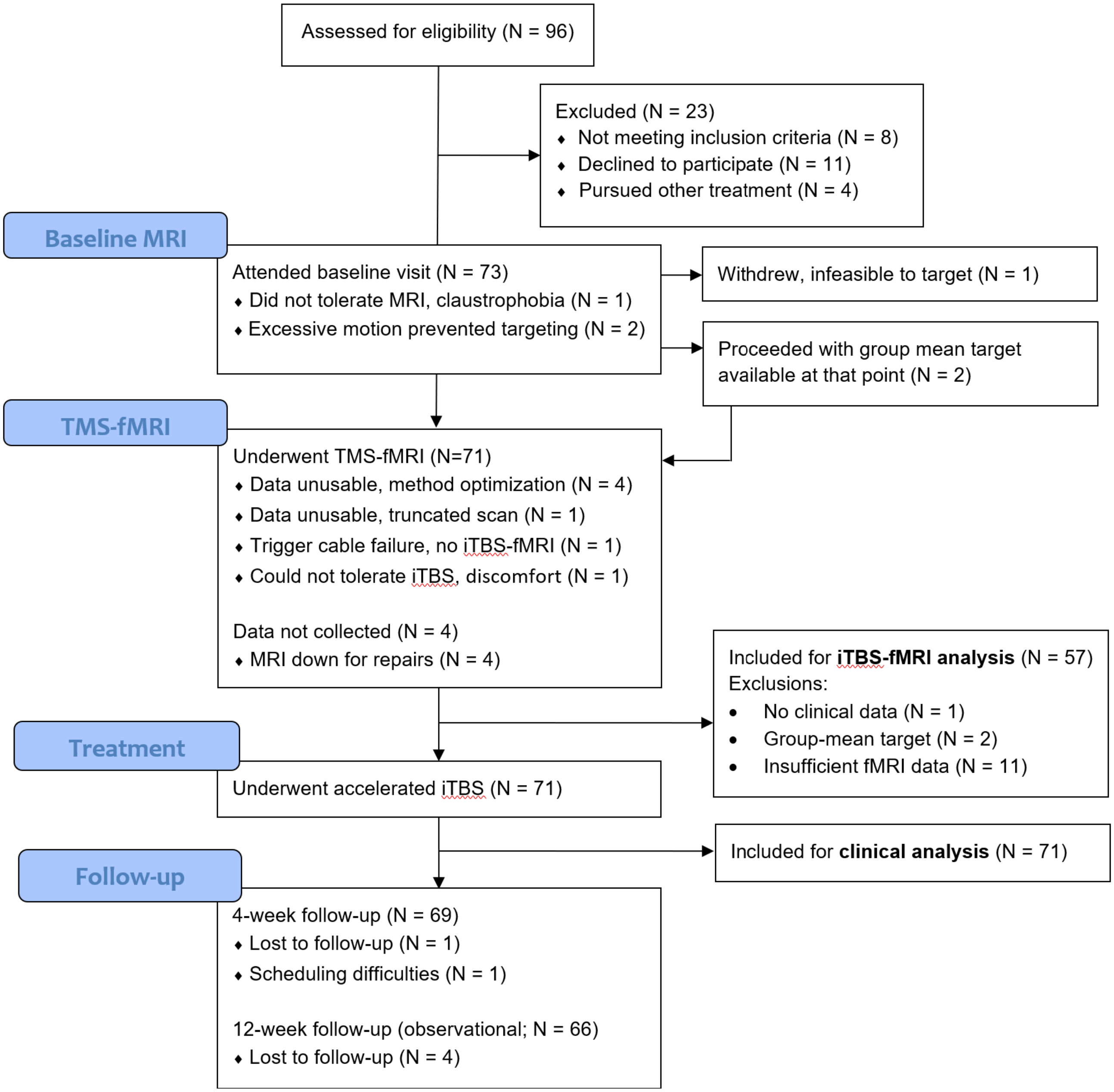


**Supplemental Figure 6. Patient flow throughout study stages and associated endpoints.** Flow of patients from first assessment, baseline, sham-controlled TMS-fMRI, open-label accelerated iTBS, and follow-up including lost data points and patients lost to follow-up. One patient withdrew at the baseline visit due to inability to tolerate the MRI, and one withdrew prior to treatment to pursue other therapies prior to treatment. Two patients had excessive motion artifacts in baseline images that were not identified at the time of scanning which precluded image-guided targeting, and were assigned group-mean targets available at that point; these individuals were included in the clinical analysis, but not the iTBS-fMRI analysis.

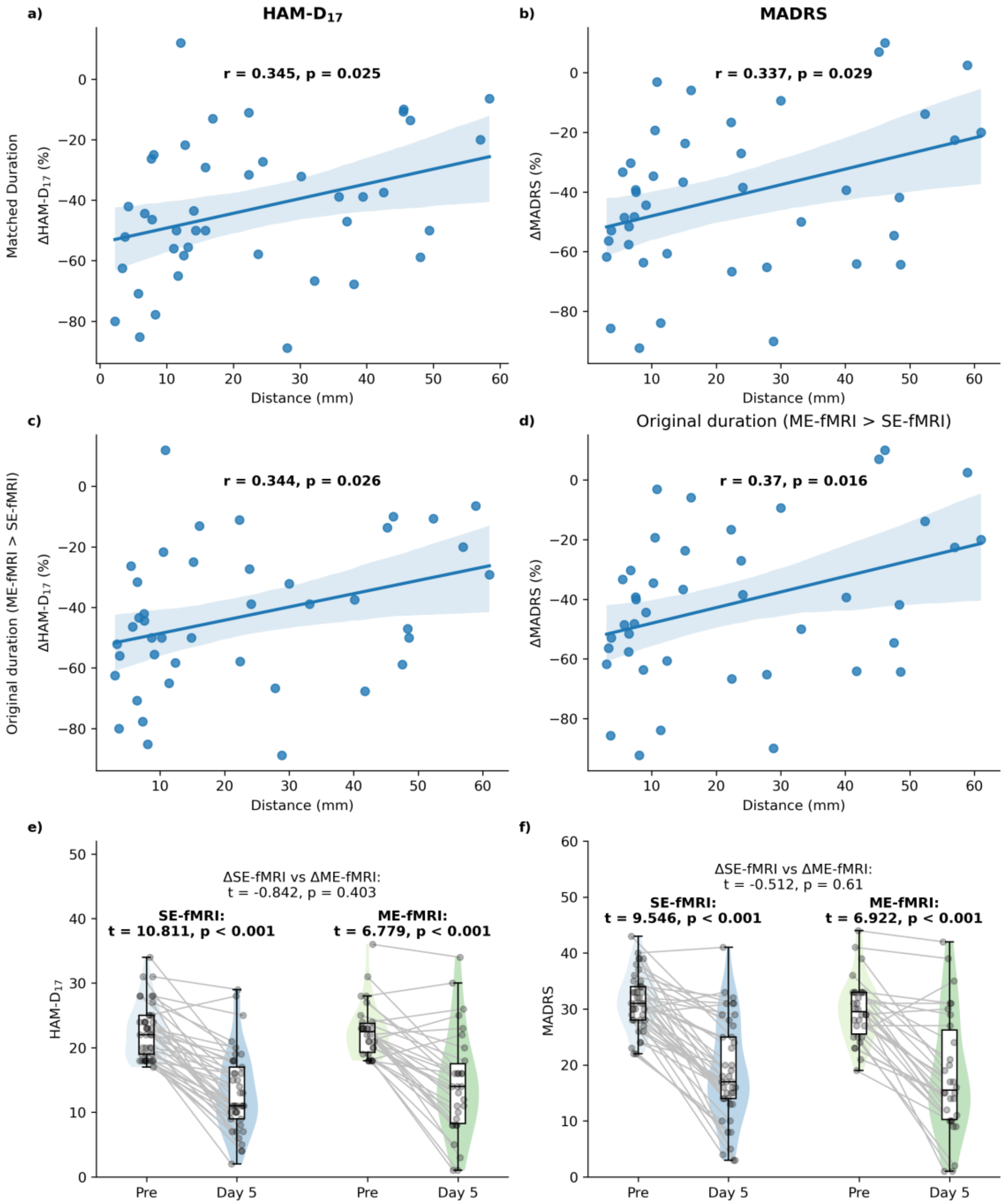


**Supplemental Figure 7. Relationship of resting state imaging protocol with clinical outcomes.** Plots show relationship of change in clinician-administered a,c) HAM-$D_{17}$ and b,d) MADRS and distance between connectivity-guided targets derived from single-echo resting-state fMRI and those derived from multi-echo resting-state fMRI scans collected at the same visit among the first N=43 patients for whom single-echo imaging was used for targeting, as well as magnitude of improvement on the HAM-$D_{17}$ and MADRS after open-label treatment. Multi-echo scans were both a,b) truncated to match the duration of single-echo scans, or c,d) with original duration retained to assess for effects of scan duration vs denoising quality. Change in e) HAM-$D_{17}$ and f) MADRS at the end of open-label treatment between patients treated using single-echo fMRI (SE-fMRI; N=43) vs multi-echo fMRI (ME-fMRI; N=28). HAM-$D_{17}$: Grid-Standardized Hamilton Depression Rating Scale, 17-item. MADRS: Montgomery-Asberg Depression Rating Scale. Bolded: $p < 0.05$.

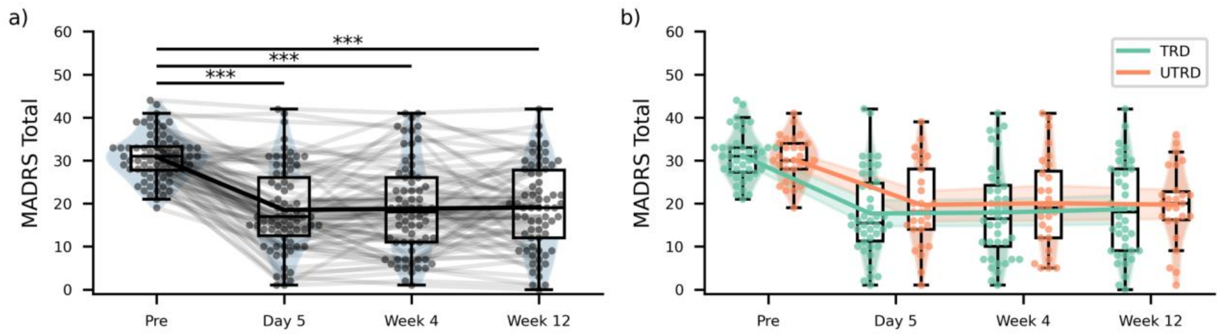


**Supplemental Figure 8. Accelerated iTBS produces rapid and sustained reductions in Montgomery-Asberg Depression Rating Scale.** Change in MADRS score, prior to rTMS treatment and again at follow-up visits. Panel (a) represents the entire cohort and panel (b) shows clinical response trajectories for TRD and UTRD groups. Bonferroni correction was applied for repeated longitudinal comparisons before significance testing. * = $p_{adj} < 0.05$, ** = $p_{adj} < 0.01$, *** = $p_{adj} < 0.001$.

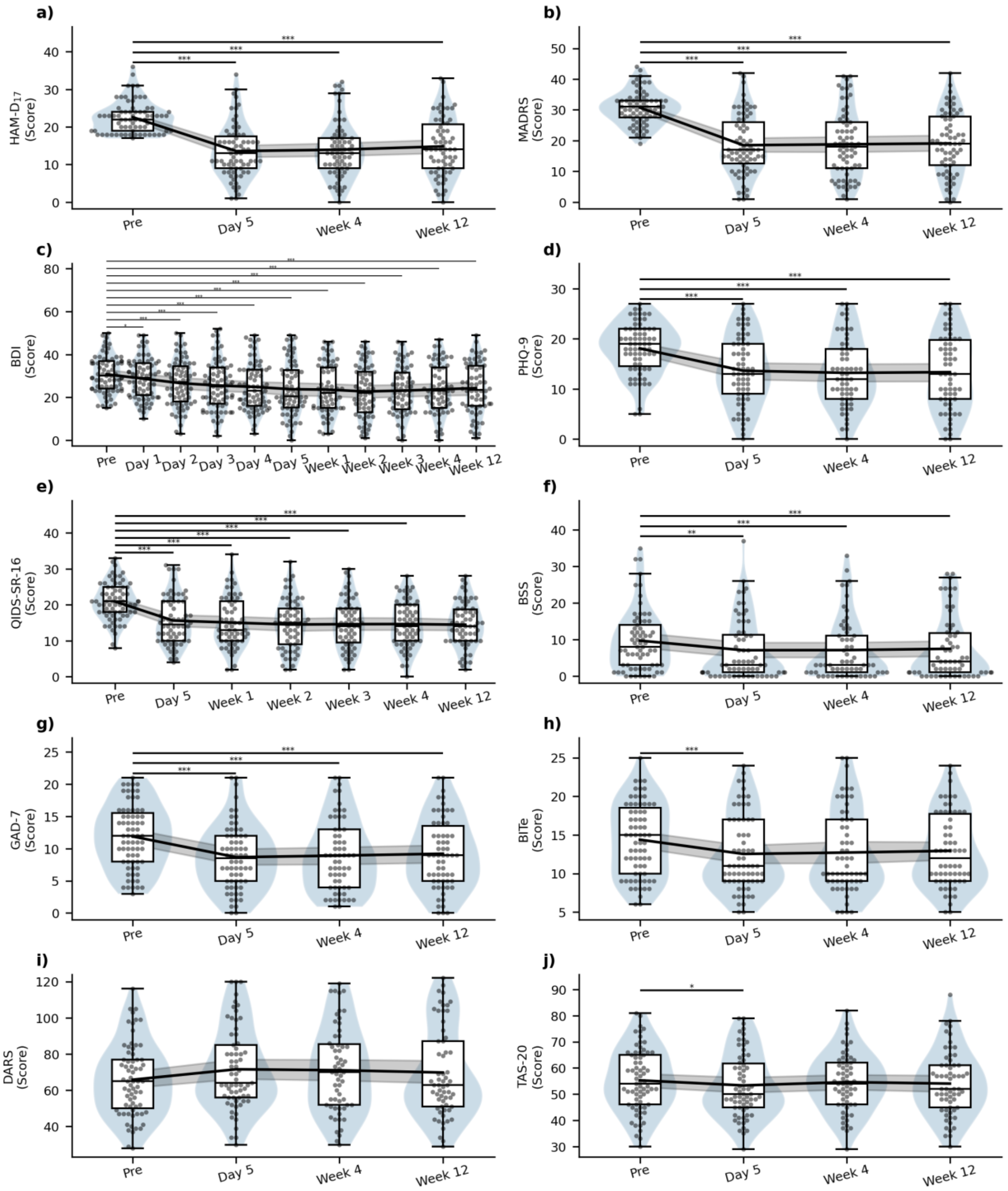


**Supplemental Figure 9.** Longitudinal change in clinician-administered clinical and self-reported symptom scales at all collected timepoints for the entire cohort. Pre and Day 5 are collected prior to and on the last day of open-label accelerated, connectivity-guided iTBS treatment. Significance of change relative to baseline is adjusted for multiple comparisons by Bonferonni correction of p-values from repeated paired t-tests. HAM-$D_{17}$: Hamilton Depression Rating Scale, 17-item. MADRS: Montgomery-Asberg Depression Rating Scale. BDI: Beck Depression Inventory. QIDS-SR-16: Quick Inventory of Depressive Symptomatology, Short Form, 16-item. PHQ-9: Patient Health Questionnaire, 9-item. BSS: Beck Scale for Suicidal Ideation. GAD-7: Generalized Anxiety Disorder scale, 7-item. BITe: Brief Irritability Test. DARS: Dimensional Anhedonia Rating Scale. TAS-20: Toronto Alexithymia Scale, 20-item. * = $p_{adj} < 0.05$. ** = $p_{adj} < 0.01$. *** = $p_{adj} < 0.001$.

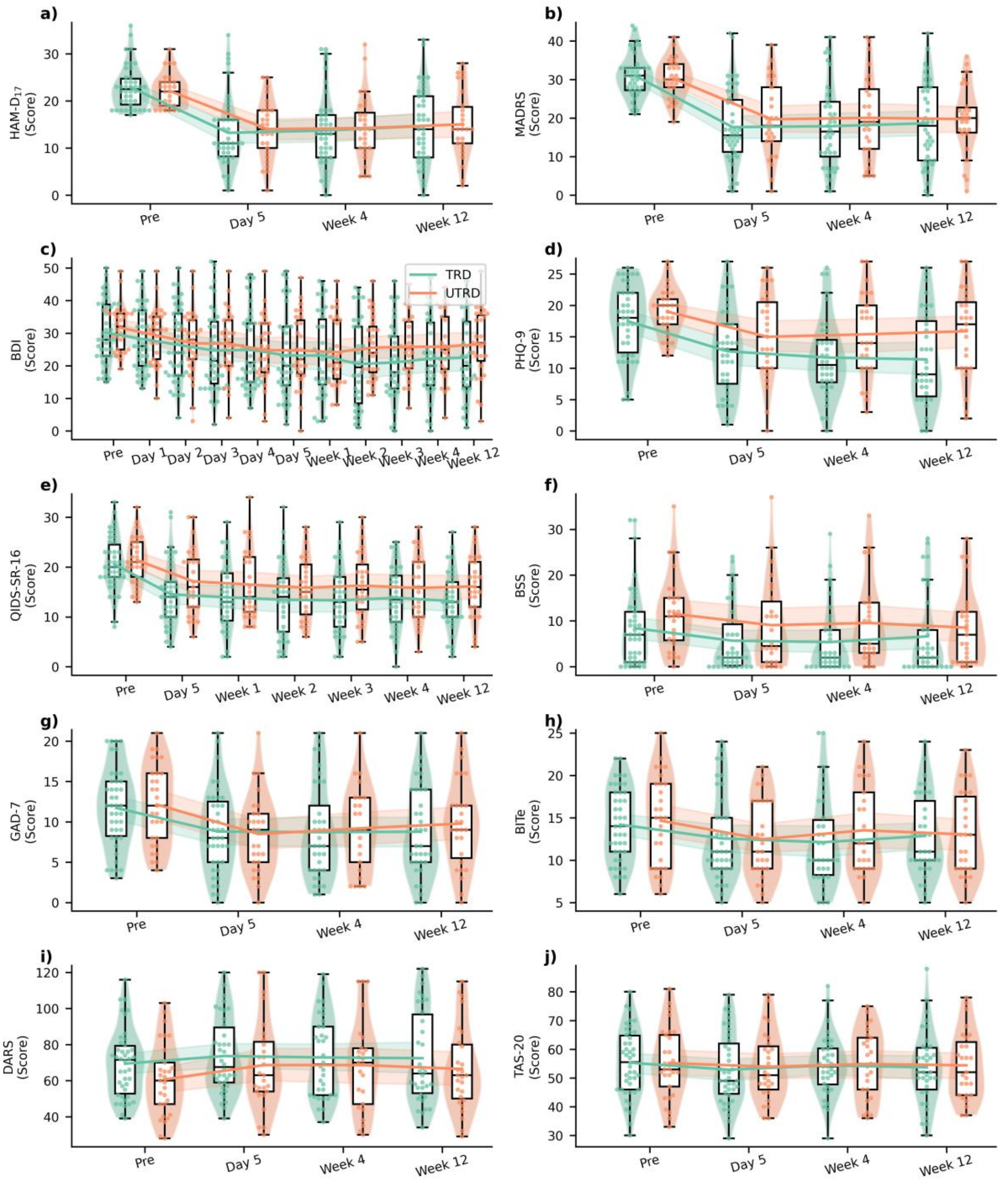


**Supplemental Figure 10.** Longitudinal change in clinician-administered clinical and self-reported symptom scales at all collected timepoints, separated by *a priori* treatment-resistance groups. Pre and Day 5 are collected prior to and on the last day of open-label accelerated, connectivity-guided iTBS treatment. Between-group differences at each timepoint were adjusted for multiple comparisons by Bonferroni correction. HAM-$D_{17}$: Hamilton Depression Rating Scale, 17-item. MADRS: Montgomery-Asberg Depression Rating Scale. BDI: Beck Depression Inventory. QIDS-SR-16: Quick Inventory of Depressive Symptomatology, Short Form, 16-item. PHQ-9: Patient Health Questionnaire, 9-item. BSS: Beck Scale for Suicidal Ideation. GAD-7: Generalized Anxiety Disorder scale, 7-item. BITe: Brief Irritability Test. DARS: Dimensional Anhedonia Rating Scale. TAS-20: Toronto Alexithymia Scale, 20-item.

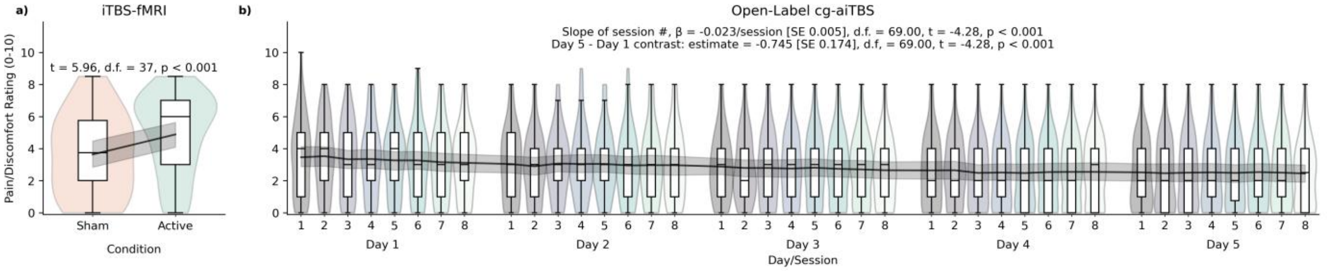


**Supplemental Figure 11.** Longitudinal change in self-reported pain and general discomfort during iTBS, both during (a) active and sham iTBS-fMRI, as well as throughout all sessions during (b) open-label accelerated treatment. a) Self-reported stimulation-associated pain and discomfort was greater during active vs sham iTBS-fMRI, as assessed by paired t-test. b) A mixed-effects model of self-reported pain/discomfort scores with a fixed and subject-level random slope of session number was fit, and used to test a contrast of self-reported pain and discomfort on the last vs first day of treatment. Over 40 sessions of open-label cg-aiTBS, self-reported pain and discomfort decreased and was significantly lower on Day 5 vs Day 1.

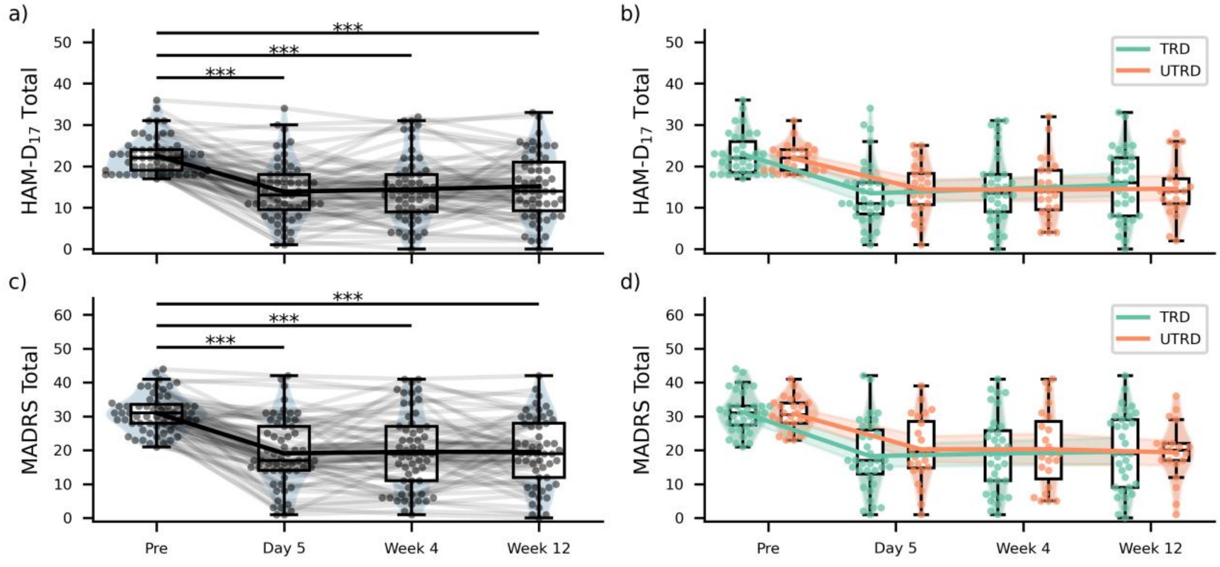


**Supplemental Figure 12. Clinician-rated outcomes in the TMS-fMRI analysis cohort.** Change in HAM-$D_{17}$ (top) and MADRS (bottom) score, prior to rTMS treatment and again at follow-up visits. Panel (a) represents the entire cohort and panel (b) shows clinical response trajectories for TRD and UTRD groups. Bonferroni correction was applied for repeated longitudinal comparisons before significance testing. * = $p_{adj} < 0.05$, ** = $p_{adj} < 0.01$, *** = $p_{adj} < 0.001$.

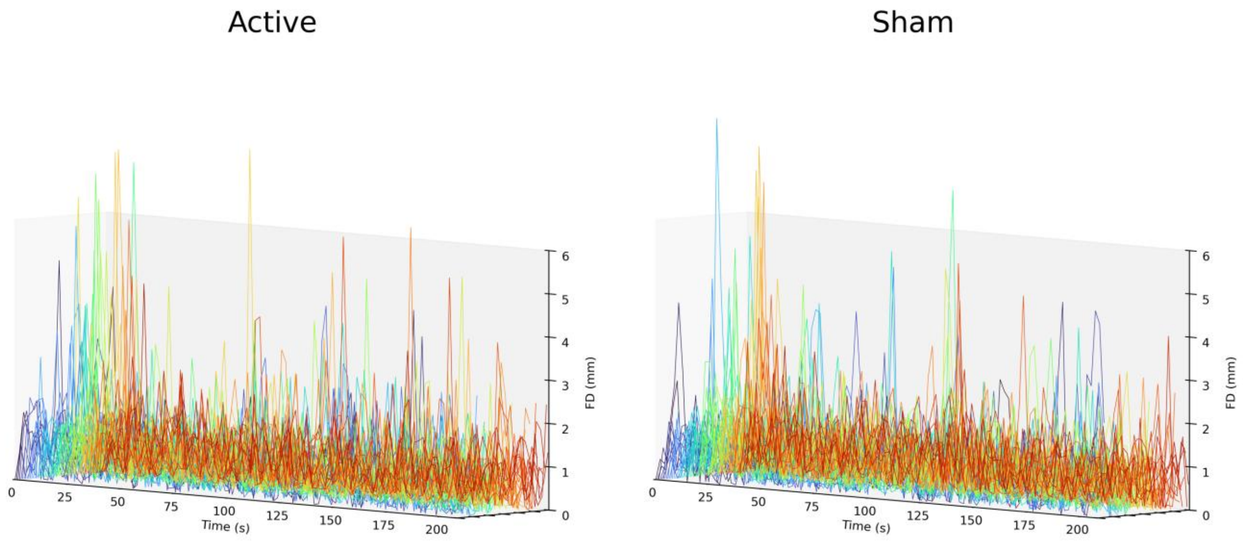


**Supplemental Figure 13. High motion during iTBS delivery and motion spikes aligned with trains.** Despite head fixation devices used during iTBS-fMRI, patients exhibited relatively high motion ($mean_{FD}$ = 0.742 +/- 0.094 mm), and trains of iTBS are associated with brief high-motion spikes, particularly the first train. Plots show individual subject-level absolute framewise displacement during iTBS-fMRI to visualize patterns of motion across the cohort in the active (left) and sham (right) condition.

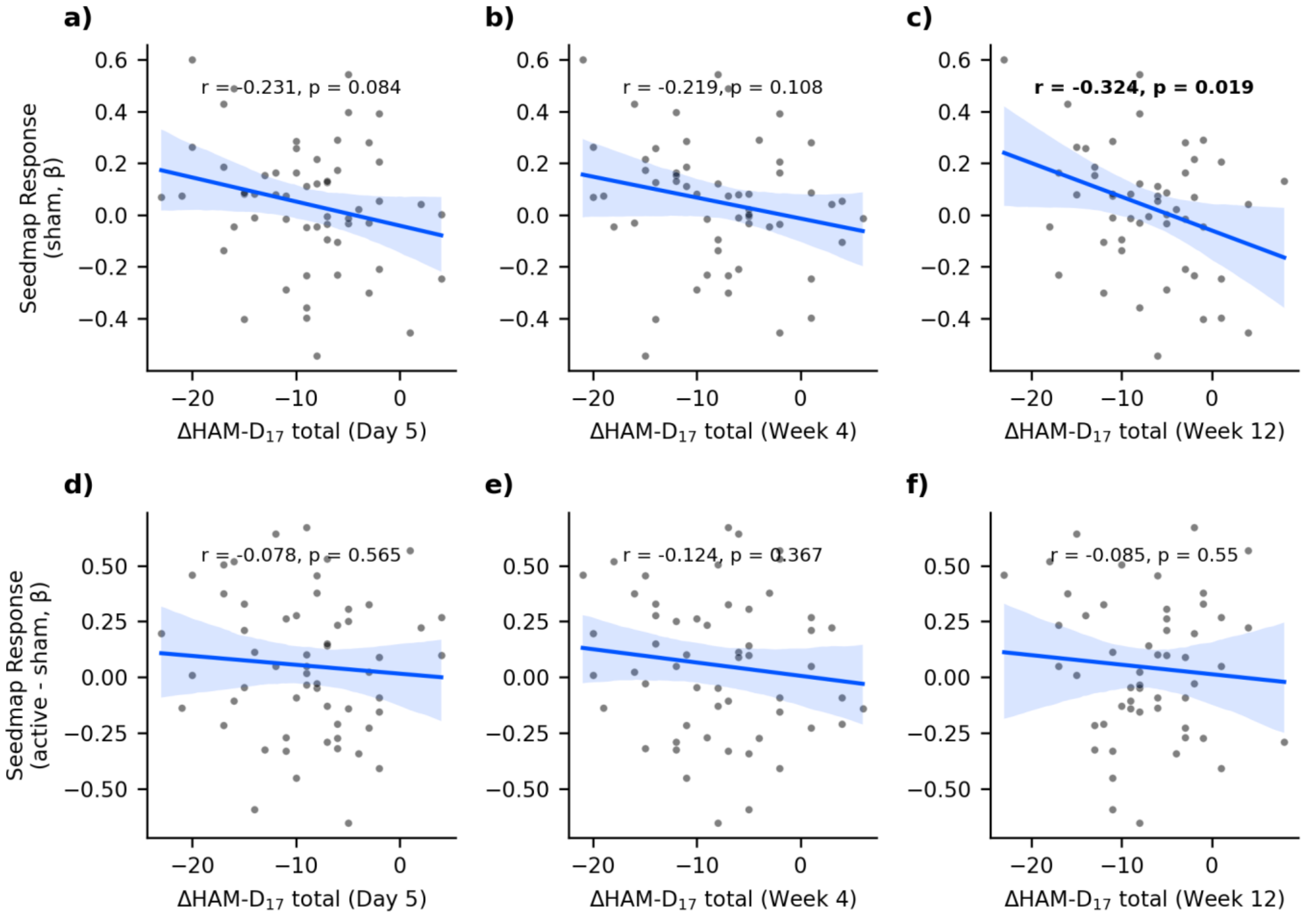


**Supplemental Figure 14. Target engagement in the sgACC-associated seedmap during sham iTBS-fMRI and the active-sham contrast is not reliably associated with clinical improvement.** Pearson's correlation of event-related responses in sgACC-associated seedmap in the sham condition (a-c) and active-sham contrast (d-f), with subsequent change in HAM-$D_{17}$ at day 5 (a,d), week 4 (b,e), and week 12 (c,f).

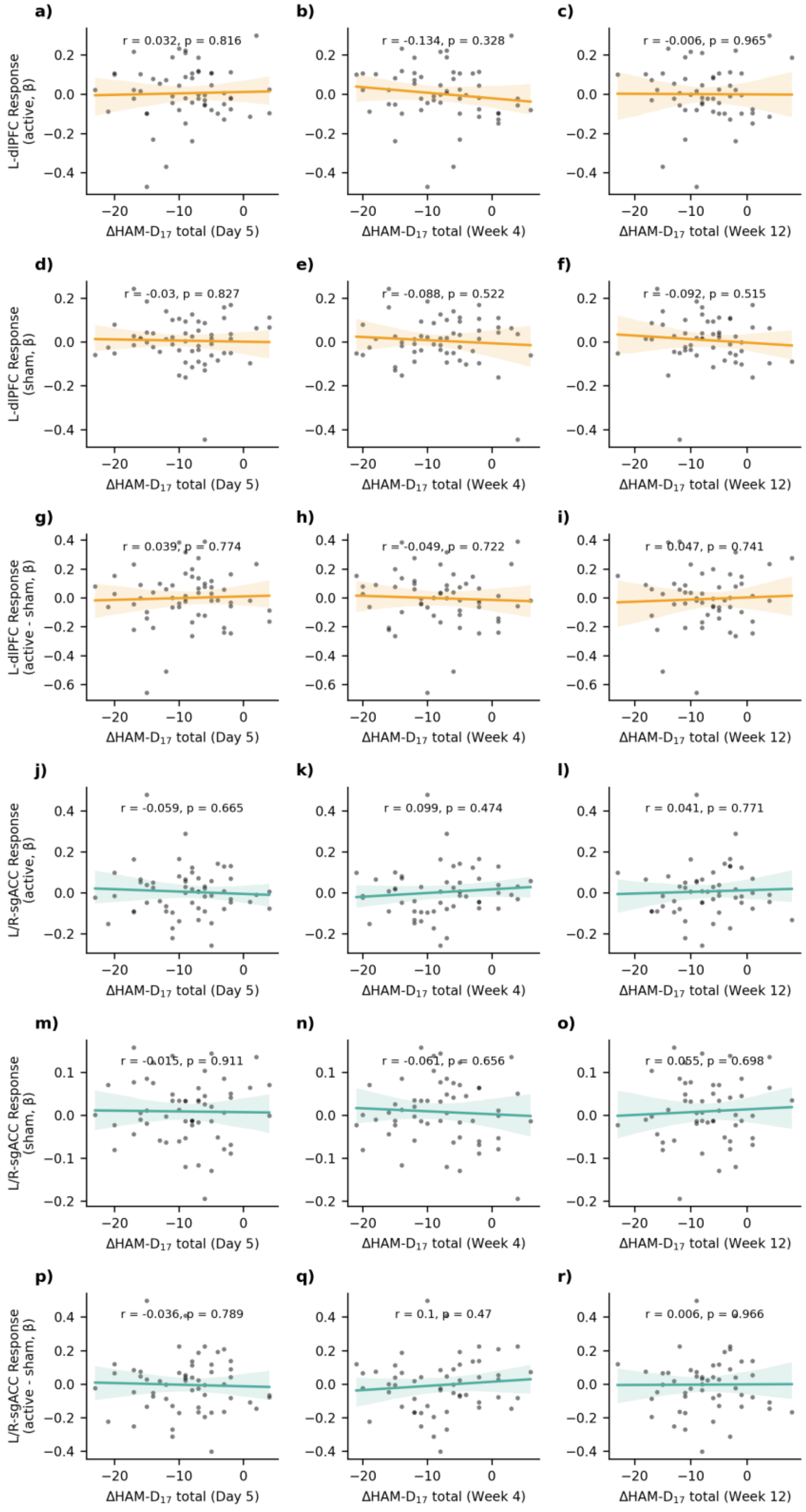


**Supplemental Figure 15. Target engagement in the left dlPFC E-field and bilateral sgACC during iTBS-fMRI are not associated with subsequent clinical improvement.** Pearson's correlation of event-related responses in each subject's left dlPFC E-field (a-i) and bilateral sgACC (j-r) in the active (a-c, j-l) and sham condition (d-f, m-o), as well as the active-sham contrast (g-l, p-r), with subsequent change in HAM-$D_{17}$ at day 5 (a,d,g,j,m,p), week 4 (b,e,h,k,n,q), and week 12 (c,f,i,l,o,r).

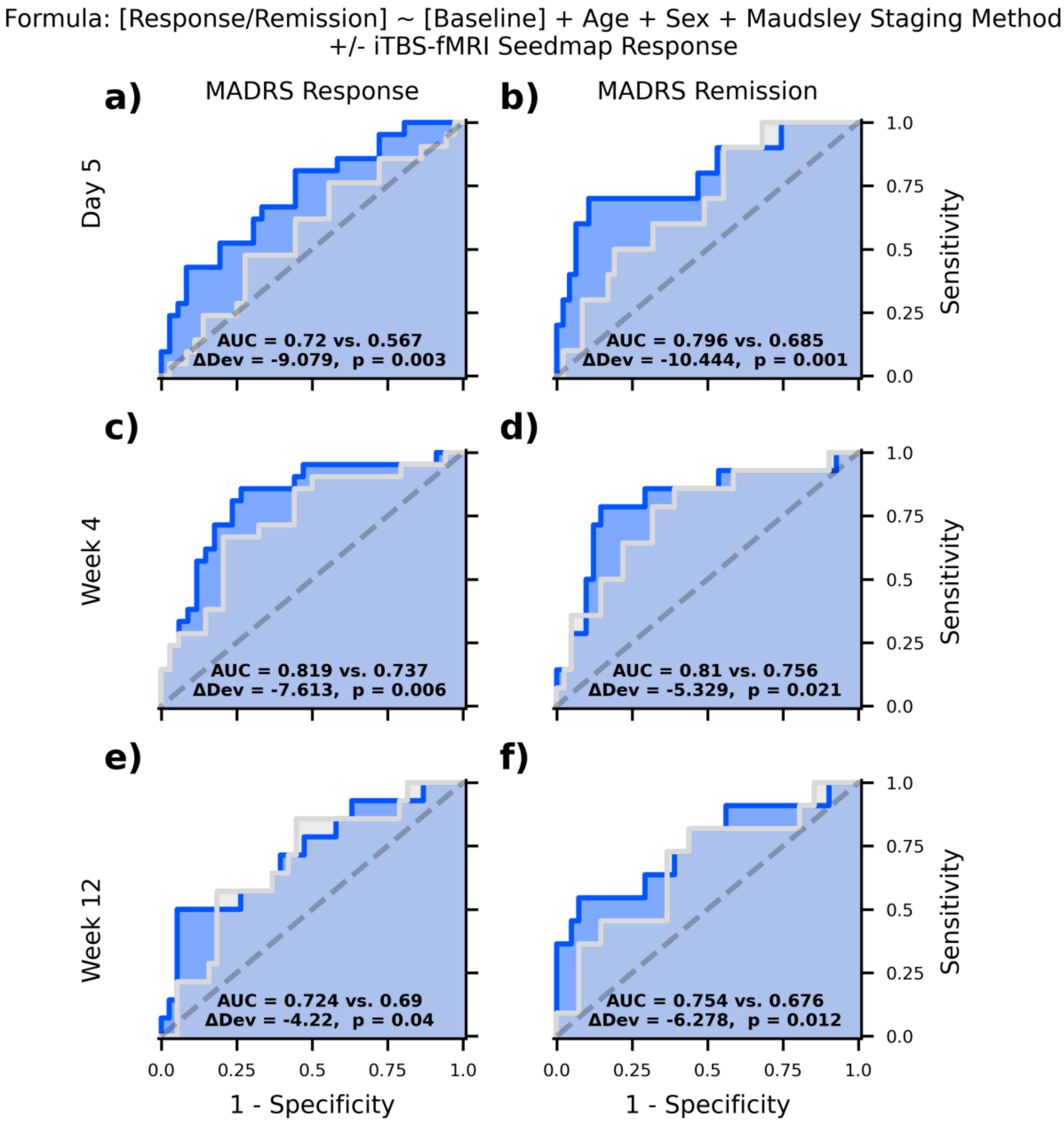


**Supplemental Figure 16. iTBS-fMRI responses in sgACC-associated seedmap predict clinical outcomes on Montgomery-Asberg Depression Rating Scale.** Receiver-operator curves demonstrating predictive capacity of nested models, testing whether logistic regression models including (blue) event-related responses during iTBS-fMRI have independent prognostic value relative to a base model (white) including demographic (age, sex) and clinical variables (baseline symptom severity, Maudsley Staging Method). Prediction of response (b,d,f) and remission (c,e,g) are shown separately at day 5 (b,c), week 4 (d,e), and week 12 (f,g) on the MADRS. AUC: area under the curve. ΔDev: change in deviance (measure of logistic regression model fit).

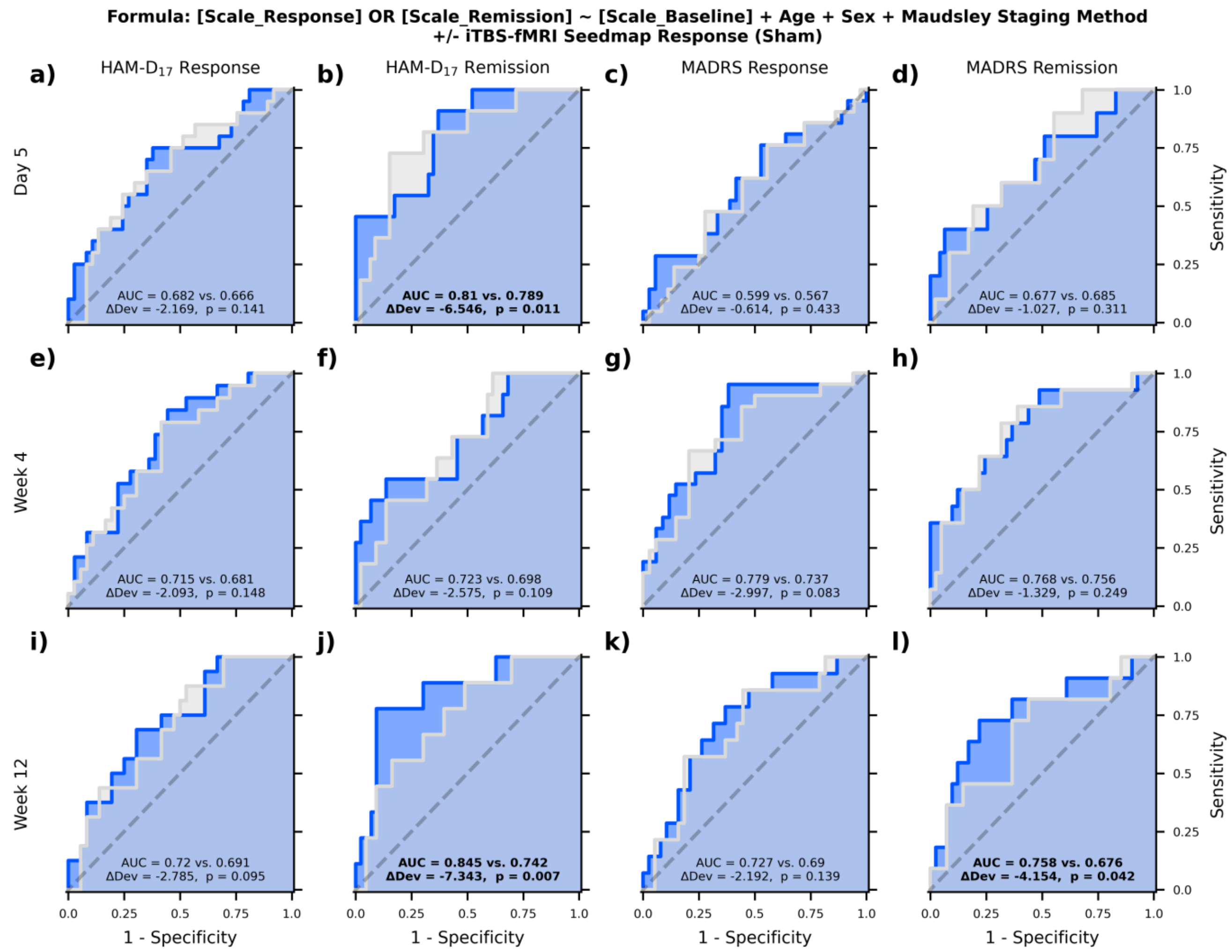


**Supplemental Figure 17. Sham iTBS-fMRI responses in the sgACC-associated seedmap do not reliably improve within-sample prediction of clinical improvement.** Receiver-operator curves demonstrating predictive capacity of nested models, testing whether logistic regression models including (blue) event-related responses during iTBS-fMRI have independent prognostic value relative to a base model (white) including demographic (age, sex) and clinical variables (baseline symptom severity, Maudsley Staging Method). Prediction of response (a,c,e,g,i,k) and remission (b,d,f,h,j,l) on the HAM-$D_{17}$ (a,b,e,f,i,j) and MADRS (c,d,g,h,k,l) are shown separately at day 5 (a-d), week 4 (e-h), and week 12 (i-l). AUC: area under the curve. ΔDev: change in deviance (measure of logistic regression model fit).

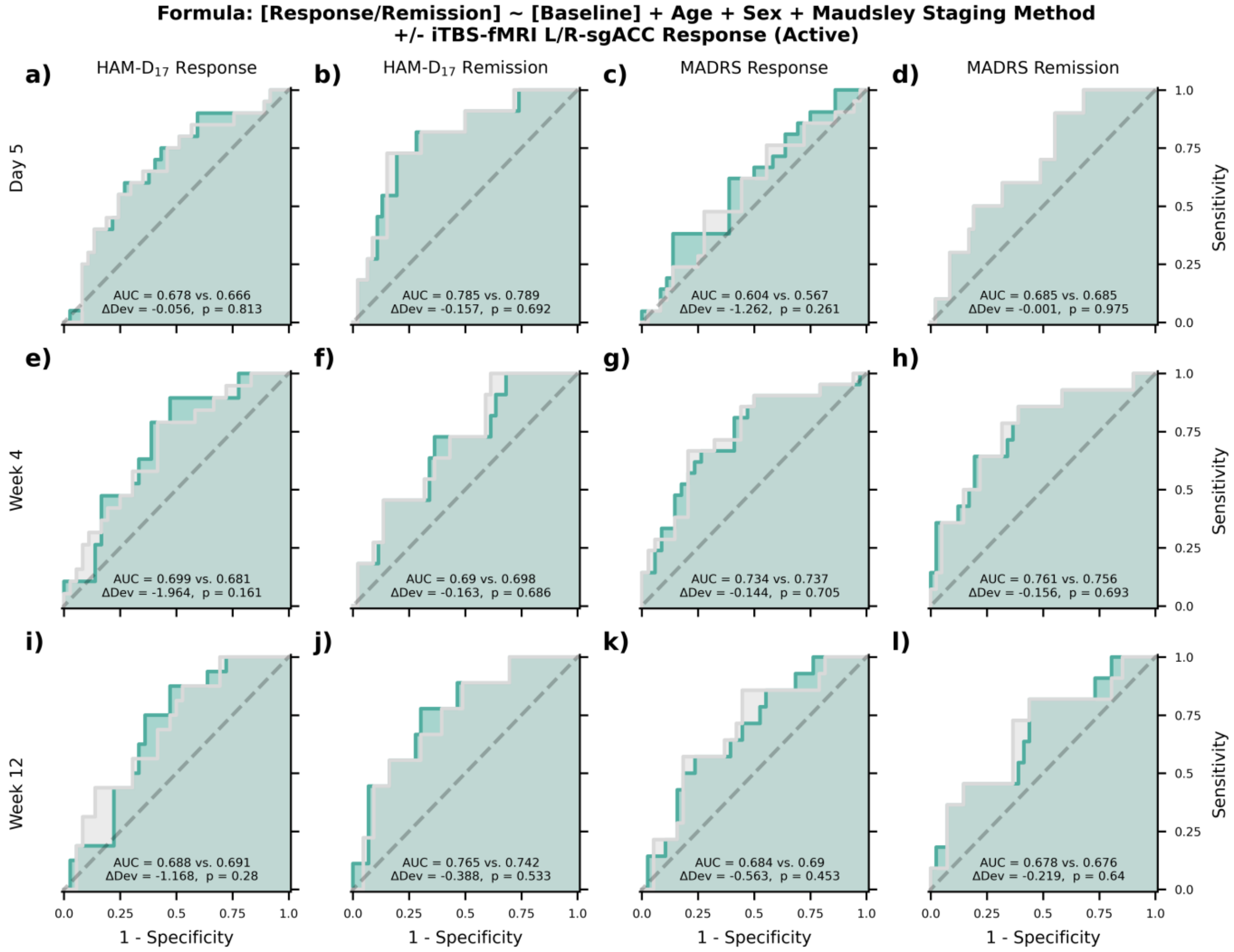


**Supplemental Figure 18. iTBS-fMRI responses in the bilateral sgACC do not improve within-sample prediction of clinical improvement.** Receiver-operator curves demonstrating predictive capacity of nested models, testing whether logistic regression models including (teal) event-related responses during iTBS-fMRI in a bilateral spherical sgACC ROI have independent prognostic value relative to a base model (white) including demographic (age, sex) and clinical variables (baseline symptom severity, Maudsley Staging Method). Prediction of response (a,c,e,g,i,k) and remission (b,d,f,h,j,l) on the HAM-$D_{17}$ (a,b,e,f,i,j) and MADRS (c,d,g,h,k,l) are shown separately at day 5 (a-d), week 4 (e-h), and week 12 (i-l). AUC: area under the curve. ΔDev: change in deviance (measure of logistic regression model fit).

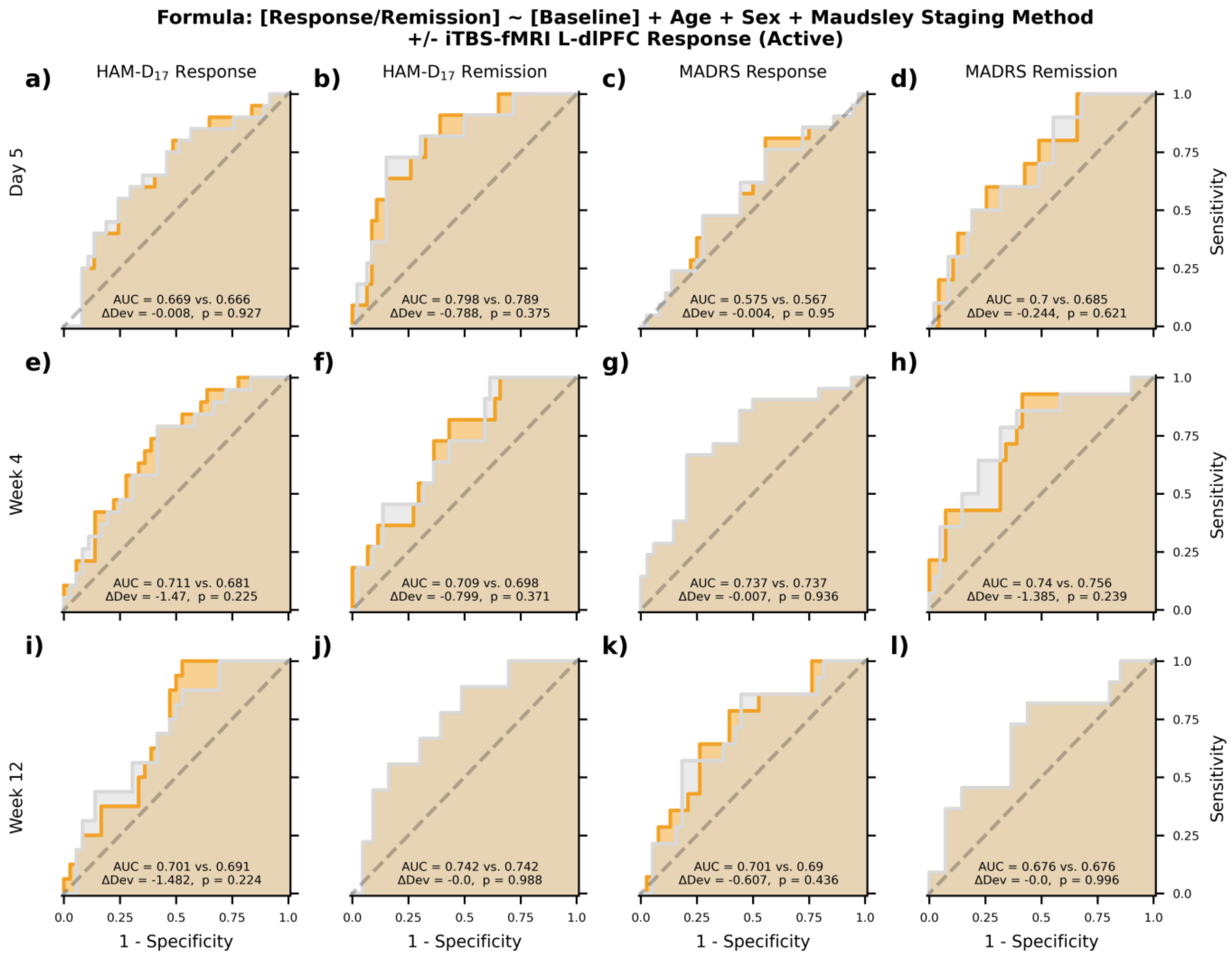


**Supplemental Figure 19. iTBS-fMRI responses in each individual's left dlPFC E-field do not improve within-sample prediction of clinical improvement.** Receiver-operator curves demonstrating predictive capacity of nested models, testing whether logistic regression models including (gold) event-related responses during iTBS-fMRI in each subject's dlPFC E-field have independent prognostic value relative to a base model (white) including demographic (age, sex) and clinical variables (baseline symptom severity, Maudsley Staging Method score). Prediction of response (a,c,e,g,i,k) and remission (b,d,f,h,j,l) on the HAM-$D_{17}$ (a,b,e,f,i,j) and MADRS (c,d,g,h,k,l) are shown separately at day 5 (a-d), week 4 (e-h), and week 12 (i-l) . AUC: ΔDev change in deviance (measure of logistic regression model fit).
