## Supplemental Methods for "Target engagement during interleaved iTBS-fMRI predicts subsequent clinical response to connectivity-guided accelerated iTBS: results of the PRISM-TRD trial"

#### *Study Design*

PRISM-TRD (predicting response via interleaved intermittent theta-burst stimulation-functional magnetic resonance imaging in treatment-resistant depression) was a pilot open-label efficacy and neuroimaging biomarker trial (NCT05813093) investigating the prognostic value of brain responses during sham-controlled iTBS-functional magnetic resonance imaging (fMRI) to predict clinical outcomes to accelerated, neuronavigated connectivity-guided intermittent theta-burst stimulation (iTBS) in patients with treatment-resistant depression (TRD). This offered two important opportunities. First, we were able to offer modern, connectivity-guided, accelerated iTBS to patients with TRD seen at our centre who may otherwise not be able to accommodate the burden of 4-6 weeks of once-daily treatment, as well as to potentially achieve clinical response in patients with active illness waitlisted for more invasive interventions (e.g. electroconvulsive therapy, focused ultrasound capsulotomy, or deep brain stimulation). Additionally, this provided us the opportunity to develop a platform for investigating mechanisms of TMS and other causally probing brain networks in patients with psychiatric illnesses. However, the neuroimaging component of this work involved significant method development and optimization with respect to MRI sequences, MRI coil configurations, TMS coil fixation, optical neuronavigation, and patient comfort as has been described by other centres developing similar research programs (1–7).

All patients underwent a baseline MRI, during which structural and fMRI data were acquired. These images were used in conjunction with electric-field modelling to identify an optimal target within the left dorsolateral prefrontal cortex (dlPFC) that was most anti-correlated with an subgenual anterior cingulate cortex (sgACC) associated seedmap. Prior to clinical treatment with an accelerated iTBS protocol, patients received one session of TMS alongside fMRI during a second MRI visit. In all cases, stimulation was delivered using neuronavigation to the personalized left dlPFC target from the first MRI visit. Thereafter, patients went on to receive 5 days of accelerated iTBS, involving 8 daily 600 pulse iTBS sessions with neuronavigation, targeting the same target and trajectory used during the TMS-fMRI scan visit.

#### *Demographic and clinical data collection*

Concomitant medications, medical and psychiatric history, as well as self-report clinical and symptom scales, were collected at baseline. The clinician administered Grid-Standardized Hamilton Depression Rating Scale (HAM-$D_{17}$)(8) and Montgomery-Åsberg Depression Rating Scale (MADRS)(9) were collected at baseline and immediately after the last treatment session on the fifth day of treatment, and at 4- and 12-week post-treatment. Maudsley Staging Method (MSM) scores were calculated from baseline HAM-$D_{17}$ total scores using suggested thresholds for severity scoring (10). Exploratory self-report scales were also acquired at baseline, the last day of treatment and at visits on weeks 4 and 12 post-treatment. These scales included measures of:

1. Global Depression: Beck Depression Inventory (BDI) (11), Patient Health Questionnaire (PHQ-9) (12), Quick Inventory of Depressive Symptomatology (QIDS-SR-16) (13)
2. Anxiety: General Anxiety Disorder, 7-item (GAD-7) (14)
3. Irritability: Brief Irritability Test (BITe) (15)
4. Anhedonia: Dimensional Anhedonia Rating Scale (DARS) (16)

5. Alexithymia: Toronto Alexithymia Scale, 20-item (TAS-20) (17)

The QIDS-SR-16 and BDI were also collected at 1-3 weeks post-treatment, and the BDI was further collected daily during open-label treatment.

Differences in clinical and demographic variables between TRD and UTRD groups were quantified using standardized mean differences (SMD). Normality of distributions was assessed by a Shapiro-Wilk's test. Continuous variables were summarized by mean and standard deviation if normally distributed, and median and interquartile range otherwise. Categorical variables were summarized by count and percent.

*Medication Management*

Prior to entering the study, patients taking benzodiazepines daily were encouraged and supported in reducing daily doses to a maximum of 1mg/day in lorazepam equivalents, and complete cessation was the goal, based on tolerability. Patients taking benzodiazepines on an as-needed basis were instructed to refrain from consuming these medications between the baseline scan (MRI visit 1) and the end of accelerated iTBS treatment. Patients were maintained on psychiatric medications at stable doses from the first visit until the post-treatment, week 4 follow-up assessment. As this trial was a real-world feasibility study of accelerated, connectivity-guided iTBS, changes to depression care, such as medication, therapy, or other neuromodulation procedures, were permitted after the 4-week post-iTBS treatment visit (i.e. during the observational phase of the study between the 4-week and the final 12-week follow-up).

*Baseline MRI acquisition*

During the baseline visit, structural MRI and resting-state functional MRI (rs-fMRI) was collected using a 3T Siemens PRISMA scanner (Siemens, Erlangen, Germany) with a standard 64-channel head/neck coil. Structural images were acquired, using a T1-weighted MPRAGE sequence (TR = 2400 ms; TE = 2.28 ms; flip angle = 8 degrees; voxel size 0.8×0.8×0.8 mm isotropic; 256mm FOV; 224 slices). To capture high-resolution images of the blood oxygenation level-dependent (BOLD) signal, we used both i) a single-echo echo-planar imaging sequence (TR = 1000ms, TE = 38.00 ms, flip angle = 60 degrees, multi-band factor = 4, voxel size = 3.0×3.0×3.0 mm isotropic, 192mm FOV, 400 volumes), and ii) a multi-echo echo-planar imaging sequence (TR = 1330 ms, TEs = 12.60/29.51/46.42/63.33ms, multi-band factor = 4, voxel size = 2.5×2.5×2.5 mm isotropic, 192mm FOV, 650 volumes). Prior to rs-MRI acquisition, participants were instructed to keep their eyes open and to let their mind wander.

*Resting-state fMRI processing and connectivity-guided target definition*

Surfaces and volumes of the T1-weighted structural images were reconstructed and parcellated using Freesurfer version 6.0.0 (18). Head meshes were reconstructed from T1 MRI for E-field simulations, using the *charm* tool from SimNIBS v4.1.0 (19). Resting-state fMRI images underwent realignment, slice timing correction, and spatial normalization with fMRIPrep v23.0.2 (20). For single-echo fMRI data, denoising used ICA-AROMA, whereas tedana was used for denoising and optimal combination of multi-echo fMRI data (21).

Personalized targets were identified based on resting-state functional MRI connectivity (rsFC) with a previously published pipeline in MATLAB (Mathworks, Massachusetts, USA) (22). First, white matter, CSF, and global signal were regressed out of the BOLD timeseries in the preprocessed rs-fMRI images, and bandpass temporal filtering was applied (0.01-0.1 Hz). Next, correlations were calculated between every brain voxel and a seed map(22). This seedmap has been reported and used previously for TMS targeting, corresponding to the sgACC as well as correlated voxels identified in subjects (N = 1000) from the Human Connectome Project dataset (22). To identify a dlPFC target most anti-correlated with this sgACC seedmap, a previously described cluster-based approach (23) was used, constrained to a dlPFC mask encompassing the anatomical dlPFC and surrounding left prefrontal regions previously reported and used to anatomically constrain TMS target selection. In cases where motion artifacts in the baseline images precluded image-guided targeting (N = 2; Suppl. Fig. 6), group mean targets available at those points were used.

SimNIBS v4.1.0 software was used to optimally derive the trajectory of the TMS coil's electric field (E-field) projected onto the left dlPFC. Briefly, E-field trajectories were sampled every 30° around points that were spatially distributed every 5mm within a 20mm radius centered on the personalized dlPFC target derived above. E-fields produced by an MRi-B91 MRI-compatible TMS coil (MagVenture A/S, Farum, Denmark) were simulated for every trajectory in this search space, using the auxiliary dipole method (19). The trajectory that maximized the predicted E-field at the target was subsequently used for all sessions where iTBS was delivered. This trajectory was used for both the interleaved TMS-fMRI (MRi-B91 coil) and open-label clinical iTBS treatments (B70 TMS coil).

*Single- and multi-echo targeting comparison*

Both single- and multi-echo resting-state fMRI were acquired in the first 43 participants for a planned methodological comparison of target localization, on the basis of previously reported advantages of multi-echo acquisition for separating BOLD from non-BOLD signal, improving resting-state connectivity estimation, and identifying TMS targets (24–26). For these participants, personalized stimulation targets used for treatment were derived from single-echo data. After completion of the first 43 participants, an exploratory interim analysis examined the spatial agreement between targets derived from the two acquisitions and its association with clinical outcome. Greater Euclidean distance between single- and multi-echo targets was associated with smaller subsequent improvement on the HAM-$D_{17}$ and MADRS. On this basis, the targeting workflow was modified prospectively so that multi-echo rs-fMRI was used to localize treatment targets for the remaining 28 participants. The remaining targeting, electric-field modeling, and treatment procedures were unchanged.

Because targeting method was changed nonrandomly during enrollment and the decision was informed partly by an exploratory association with clinical outcomes, analyses comparing participants targeted with single- versus multi-echo rs-fMRI were considered exploratory and were not interpreted as tests of the comparative efficacy of the two methods.

*Interleaved TMS-fMRI*

The interleaved TMS-fMRI setup was composed of a unique combination of commercially available devices. Initially, an MRi-B91 air-cooled MR-compatible TMS coil (Magventure, Denmark) was integrated with the posterior shell of a 20-channel 3T head/neck coil (Siemens, Germany), in addition to a 4-channel flexible MR receiver body coil (Siemens, Germany) wrapped around the participants head and the TMS coil. We later transitioned to using the posterior shell of a 64-channel 3T head/neck coil with the further addition of a 4-channel carotid coil (Siemens,

Germany) on the right side of the head opposite the rTMS coil. These changes were implemented to improve temporal signal-to-noise ratio (tSNR), particularly in deep brain regions such as the sgACC.

In all cases, prior to placing the flexible body coil over the participant's head, the TMS coil was positioned over the personalized target site in the dlPFC, guided by optical neuronavigation (Brainsight, Rogue Research; Polaris Vega ST, Northern Digital Inc.). The MRI-B91 coil was stabilized with an MR-compatible coil holder (MRI-Coil Holder, part 9016B0471; Magventure, Denmark) to ensure precise placement throughout the scanning session. Throughout, the patient's head laid in a vacuum head cushion (Siemens, Germany; 04765462) to which suction was applied shortly before the flexible body coil was placed, improving fixation of head position and providing additional physical support of the coil while scanning.

Interleaved iTBS-fMRI was performed using the FDA/Health Canada approved iTBS pulse pattern which comprised 2-second pulse trains with 10 bursts (each burst = 3 pulses at 50 Hz; bursts repeated at 5 Hz), followed by an 8-second inter-train interval (6, 27, 28). The stimulation was delivered at 100% of the resting motor threshold (rMT). Stimulation was interleaved with an fMRI acquisition, using a multi-band EPI sequence (TR = 2000 ms, TE = 38 ms, voxel size = 3.0×3.0×3.0 mm, 192mm FOV, 40 slices, 20% slice gap, multiband factor = 4). For sham stimulation, a 1 cm thick polylactic acid block was placed outside of the participant's field of view, between the scalp and the TMS coil. The block minimized cortical stimulation, while preserving scalp sensations. This setup ensured a controlled comparison between active and sham conditions, mitigating the confounding effects of sensory stimuli and discomfort/pain. Participants were asked to report any side effects, and guess the order in which they received active or sham stimulation. Answers were recorded to assess the integrity of the sham. After subjective reports that iTBS-fMRI was associated with greater discomfort than open-label iTBS among the first 28 patients, we subsequently began to systematically ask patients to rate the level of discomfort they experienced during each stimulation condition.

*iTBS-fMRI data processing*

Data acquired during iTBS-fMRI were preprocessed with *fMRIprep* 25.2.5 (20). Preprocessed fMRI images were manually reviewed for quality. As susceptibility distortion correction (SDC) introduced inappropriate distortions of anatomy local to the ventromedial prefrontal cortex in a subset of patients with signal dropout in this region, SDC was disabled. Similarly, functional space brain masks derived from the bold reference were found to exclude areas of the ventromedial prefrontal cortex and sgACC in some subjects experiencing signal dropout, so anatomical space masks were transformed to functional space and used in place of masks generated from the bold reference in fMRIprep's BOLD preprocessing workflows.

Given the high motion interspersed with stimulus-locked spikes and poor temporal signal-to-noise ratio (tSNR) observed in this dataset, ICA-AROMA was run using *fmripost-aroma* to provide confound regressors for non-aggressive ICA-based denoising based on empirical data supporting its efficacy in high motion datasets (29–32). Subsequently, whole-brain voxelwise first-level regressions were performed with an event-related iTBS regressor, 12-parameter motion model (X/Y/Z translation and rotation, plus first temporal derivatives), CSF and white matter (plus their first derivatives), non-aggressive ICA-AROMA regressors for each subject, with voxelwise AR(1) prewhitening. Additionally, motion spikes were removed with aggressive volume censoring, including all volumes where framewise displacement exceeded 1.75mm as well as one volume before and 4 volumes, to address long-lasting (~10 sec) spin-history effects of large motion spikes (33). All denoising and event-related response estimation was performed in a single regression using fMRIprep's minimally-preprocessed BOLD timeseries. First-level betas (active, sham) or their contrast (active-sham) were used in subsequent second-level analyses.

Activation in regions-of-interest (ROIs) was assessed by averaging first-level voxelwise betas, or their contrast, within masks or seedmaps. The bilateral spherical sgACC mask was generated at the same target coordinates and size (10mm radius, MNI6 ±6, 16, 10) as previously described (23). Left dlPFC E-field ROIs were generated by robust thresholding of E-field volumes generated by SimNIBS at 90% of the 99.9$^{th}$ percentile of the whole image (i.e. top 10%, robust to outliers) to address variable ROI sizes when using a naïve top 10% threshold driven by a very small number of superficial voxels with high E-fields in a subset of patients. Seedmap activation was extracted as a weighted average of voxels based on their beta weights in the seedmap.

*Intermittent theta-burst stimulation intervention*

This trial utilized the Health Canada/FDA-approved 600-pulse iTBS protocol, comprised of 2-second pulse trains with 10 bursts (3 pulses each, at 50 Hz; bursts repeated at 5 Hz), followed by an 8-second inter-train interval(28). With a total of 20 trains, this stimulation protocol can be completed in 3 minutes and 20 seconds. 600-pulse iTBS was used based on its established clinical efficacy (28), lack of reliable differences in neurophysiological (34, 35) effects of 1800-pulse iTBS. On the first day of treatment, the rMT was determined using the treatment coil. The first treatment stimulation was administered at 100% rMT and increased to 110% rMT over the first three treatments.

The intervention was delivered repeatedly over 5-days, with eight 600-pulse iTBS sessions administered daily with a 50-minute intersession interval (ISI). Each treatment was delivered under neuronavigated guidance, with trackers affixed and calibration performed before each session. Adverse events, as well as pain and discomfort (as a single measure), were assessed after each treatment session.

*Statistical Analysis*

Longitudinal change and within-subject between conditions differences (sham and active) were tested by two-sided paired *t*-tests. Between-group (TRD vs UTRD) differences at timepoints were assessed by a two-sided independent *t*-test. One-sample or paired t-tests (both two-sided), respectively, were used to test activation in ROIs. Pearson's correlation coefficient was used to assess correlation between change in HAM-$D_{17}$ at the end of treatment and ROI activation. These primary outcomes were not adjusted for multiple comparisons.

Contrasts of estimated marginal means from mixed-effects models tested overall treatment effects over immediate post, week 4, and week 12 timepoints vs pre. Specifically, all mixed-effects models testing clinical outcomes and relationships thereof included a fixed-effect multi-level categorical effect of time (pre, post, week 4, week 12) and binary categorical (pre vs post-treatment) random effect nested in subject as well as a subject-level random intercept. A subject-level effect of time was included in these models to align them with subsequent models used for testing cross-level interactions between time and iTBS-fMRI outcomes. In all mixed-effects models, degrees of freedom were estimated by the Kenward-Roger method. A contrast vector of (-1,1/3,1/3,1/3) was applied across pre, post, week 4, and week 12 timepoints. Significance of overall treatment effects were adjusted for multiple comparisons by Bonferroni correction, although the HAM-$D_{17}$ is reported uncorrected as the primary clinical outcome in text. Post-hoc tests of longitudinal within- or between-subject differences used paired or independent (respectively) t-tests (both two-tailed) adjusted for multiple comparisons by Bonferroni correction. Relationship of covariates, such as iTBS-fMRI ROI responses, with clinical improvement in across all timepoints was assessed using the same contrast in similar-mixed effects models including an interaction with the fixed effect of time. To aid interpretation of the strength of relationships, clinical outcome measures and brain responses both z-scored such that the interaction term would represent a standardized association. For each ROI/condition, relationships between measured iTBS-fMRI response and clinical improvement on each scale was adjusted for multiple comparison by the two-stage Benjamini-Hochberg method (36).

For prognostic models assessing the predictive value of demographic, clinical, and neuroimaging variables, logistic regression models of response and remission, measured by defined cut-off scores (HAM-$D_{17}$ and MADRS), were fitted. These models were then used to create receiver operating characteristic (ROC) curves, demonstrating trade-offs in model performance in terms of sensitivity and specificity across a range of cut-off thresholds. The fit of nested models to the data was compared using analysis of deviance. Specifically, this test evaluated the improvement in prognostic power resulting from the addition of neuroimaging variables (dlPFC-sgACC rsFC or iTBS-fMRI sgACC deactivation) to a base model including the following demographic and clinical variables: age, sex, baseline depression severity (on the same scale used to measure the relevant outcome), and treatment resistance (Maudsley Staging Method score). ROC analyses were conducted using the pROC and precrec packages in R (R Laboratories, Vienna).

The interim analysis which motivated transition to multi-echo rs-fMRI for the targeting pipeline in the remainder of the cohort compared targets derived from fMRI timeseries denoised with ICA-AROMA to the optimally-combined and denoised timeseries produced by tedana (as described above). Specifically, change in clinician-administered HAM-$D_{17}$ rating scale was correlated with Euclidean distance between the 'actual' single-echo fMRI-derived target, and the theoretical alternative target derived from the 'unused' multi-echo fMRI timeseries and target. As the multi-echo sequence we collected was longer than the single-echo scan, this analysis was repeated separately using both multi-echo scans truncated to match the duration of single-echo scans, as well as the original untruncated scans of differing durations. A negative correlation in this case would suggest that greater distance/dissimilarity predicts lower improvement (inferiority of single echo vs multi-echo), positive correlation would suggest greater distance predicts greater improvement (superiority of single echo vs multi-echo), or weak/no correlation (non-inferiority, no benefit). To retrospectively test whether efficacy differed by targeting method, an independent t-test of difference in change in HAM-$D_{17}$ between the two groups was performed. Post-hoc within-group paired t-tests are presented for descriptive statistics, but are not corrected for multiple comparison.

In all analyses, a p-value threshold of $\alpha = 0.05$ was used to assess significance. Univariate tests were performed in Python using the *numpy* and *scipy* packages, and correction of p-values for multiple comparisons in longitudinal analyses was performed using the *statsmodels* package. Statistical plots were generated using *matplotlib* and *seaborn*. Multivariable logistic and linear mixed-effects models were fit in R. These analyses used the standard *stats* library as well as the *lme4*, *lmerTest*, and *pbkrtest* packages. The study overview figure was created in BioRender. Group-mean baseline functional connectivity and target locations were visualized using *nilearn*.

Example subject-level connectivity and target location was captured from the Brainsight neuronavigation software.

### References


1. Woolgar A, Feredoes E, Assem M, et al.: Consensus guidelines for the use of concurrent TMS-fMRI in cognitive and clinical neuroscience. Nat Protoc 2025; 1–17

2. Navarro de Lara LI, Windischberger C, Kuehne A, et al.: A novel coil array for combined TMS/fMRI experiments at 3 T. Magnetic Resonance in Medicine 2015; 74:1492–1501

3. Navarro De Lara LI, Tik M, Woletz M, et al.: High-sensitivity TMS/fMRI of the Human Motor Cortex Using a Dedicated Multichannel MR Coil. NeuroImage 2017; 150:262–269

4. Ge R, Humaira A, Gregory E, et al.: Predictive Value of Acute Neuroplastic Response to rTMS in Treatment Outcome in Depression: A Concurrent TMS-fMRI Trial. AJP 2022; 179:500–508

5. Bossus L, Dickson J, Blaine C, et al.: Causal connectivity maps derived from single-pulse interleaved TMS/fMRI. Sci Rep 2026; 16:3070

6. Chang K-Y, Tik M, Mizutani-Tiebel Y, et al.: Neural response during prefrontal theta burst stimulation: Interleaved TMS-fMRI of full iTBS protocols. NeuroImage 2024; 291:120596

7. Xiong Y, Burke M, Melo L, et al.: Optimizing Network-Level TMS-fMRI: Benchmarking a Novel TMS-Compatible “Sushi” MR Coil [Internet]2026; 2026.01.27.701271[cited 2026 Feb 27] Available from: https://www.biorxiv.org/content/10.64898/2026.01.27.701271v1

8. Williams JBW, Kobak KA, Bech P, et al.: The GRID-HAMD: standardization of the Hamilton Depression Rating Scale. International Clinical Psychopharmacology 2008; 23:120

9. Montgomery SA, Åsberg M: A New Depression Scale Designed to be Sensitive to Change. Br J Psychiatry 1979; 134:382–389

10. Fekadu A, Donocik JG, Cleare AJ: Standardisation framework for the Maudsley staging method for treatment resistance in depression. BMC Psychiatry 2018; 18:100

11. Beck AT, Ward CH, Mendelson M, et al.: An inventory for measuring depression. Arch Gen Psychiatry 1961; 4:561–571

12. Kroenke K, Spitzer RL, Williams JB: The PHQ-9: validity of a brief depression severity measure. J Gen Intern Med 2001; 16:606–613

13. Rush AJ, Trivedi MH, Ibrahim HM, et al.: The 16-Item quick inventory of depressive symptomatology (QIDS), clinician rating (QIDS-C), and self-report (QIDS-SR): a psychometric evaluation in patients with chronic major depression. Biological Psychiatry 2003; 54:573–583

14. Spitzer RL, Kroenke K, Williams JBW, et al.: A brief measure for assessing generalized anxiety disorder: the GAD-7. Arch Intern Med 2006; 166:1092–1097

15. Holtzman S, O’Connor BP, Barata PC, et al.: The Brief Irritability Test (BITe): A Measure of Irritability for Use Among Men and Women. Assessment 2015; 22:101–115

16. Rizvi SJ, Quilty LC, Sproule BA, et al.: Development and validation of the Dimensional Anhedonia Rating Scale (DARS) in a community sample and individuals with major depression. Psychiatry Res 2015; 229:109–119

17. Bagby RM, Parker JD, Taylor GJ: The twenty-item Toronto Alexithymia Scale--I. Item selection and cross-validation of the factor structure. J Psychosom Res 1994; 38:23–32

18. Fischl B: FreeSurfer. NeuroImage 2012; 62:774–781

19. Gomez LJ, Dannhauer M, Peterchev AV: Fast computational optimization of TMS coil placement for individualized electric field targeting. NeuroImage 2021; 228:117696

20. Esteban O, Markiewicz CJ, Blair RW, et al.: fMRIPrep: a robust preprocessing pipeline for functional MRI. Nat Methods 2019; 16:111–116

21. DuPre E, Salo T, Ahmed Z, et al.: TE-dependent analysis of multi-echo fMRI with tedana. JOSS 2021; 6:3669

22. Cash RFH, Cocchi L, Lv J, et al.: Personalized connectivity-guided DLPFC-TMS for depression: Advancing computational feasibility, precision and reproducibility. Human Brain Mapping 2021; 42:4155–4172

23. Fox MD, Buckner RL, White MP, et al.: Efficacy of Transcranial Magnetic Stimulation Targets for Depression Is Related to Intrinsic Functional Connectivity with the Subgenual Cingulate. Biological Psychiatry 2012; 72:595–603

24. Lynch CJ, Power JD, Scult MA, et al.: Rapid Precision Functional Mapping of Individuals Using Multi-Echo fMRI. Cell Reports 2020; 33:108540

25. Lynch CJ, Elbau I, Liston C: Improving precision functional mapping routines with multi-echo fMRI. Current Opinion in Behavioral Sciences 2021; 40:113–119

26. Kundu P, Voon V, Balchandani P, et al.: Multi-echo fMRI: A review of applications in fMRI denoising and analysis of BOLD signals. NeuroImage 2017; 154:59–80

27. Huang Y-Z, Chen R-S, Rothwell JC, et al.: The after-effect of human theta burst stimulation is NMDA receptor dependent. Clin Neurophysiol 2007; 118:1028–1032

28. Blumberger DM, Vila-Rodriguez F, Thorpe KE, et al.: Effectiveness of theta burst versus high-frequency repetitive transcranial magnetic stimulation in patients with depression (THREE-D): a randomised non-inferiority trial. The Lancet 2018; 391:1683–1692

29. Pruim RHR, Mennes M, van Rooij D, et al.: ICA-AROMA: A robust ICA-based strategy for removing motion artifacts from fMRI data. Neuroimage 2015; 112:267–277

30. Pruim RHR, Mennes M, Buitelaar JK, et al.: Evaluation of ICA-AROMA and alternative strategies for motion artifact removal in resting state fMRI. NeuroImage 2015; 112:278–287

31. De Blasi B, Caciagli L, Storti SF, et al.: Noise removal in resting-state and task fMRI: functional connectivity and activation maps. J Neural Eng 2020; 17:046040

32. Ciric R, Wolf DH, Power JD, et al.: Benchmarking of participant-level confound regression strategies for the control of motion artifact in studies of functional connectivity. NeuroImage 2017; 154:174–187

33. Power JD, Mitra A, Laumann TO, et al.: Methods to detect, characterize, and remove motion artifact in resting state fMRI. NeuroImage 2014; 84:320–341

34. McCalley DM, Lench DH, Doolittle JD, et al.: Determining the optimal pulse number for theta burst induced change in cortical excitability. Sci Rep 2021; 11:8726

35. Desforges M, Hadas I, Mihov B, et al.: Dose-response of intermittent theta burst stimulation of the prefrontal cortex: A TMS-EEG study. Clinical Neurophysiology 2022; 136:158–172

36. Benjamini Y, Krieger AM, Yekutieli D: Adaptive linear step-up procedures that control the false discovery rate. Biometrika 2006; 93:491–507
