## Supplemental Results for "Target engagement during interleaved iTBS-fMRI predicts subsequent clinical response to connectivity-guided accelerated iTBS: results of the PRISM-TRD trial"

*Single- versus multi-echo resting-state fMRI targeting*

An exploratory interim analysis was conducted after the first 43 participants, for whom both single- and multi-echo resting-state fMRI were acquired and single-echo data were used for target localization. Here, greater spatial divergence between targets derived from single- and multi-echo data was associated with less subsequent clinical improvement in patients where target localization used single-echo data (HAM-$D_{17}$: r = 0.344, p = 0.026; MADRS: r = 0.370, p = 0.016; Suppl. Fig. 7a–d). Similarly, when multi-echo acquisition duration was truncated to match single-echo scans, target distance was still associated with less improvement on the HAM-$D_{17}$ (r = 0.345, p = 0.025) and MADRS (r = 0.337, p = 0.029). Consequently, multi-echo data were subsequently used for target localization in the remaining 28 participants. Clinical outcomes did not differ significantly between participants targeted using single- versus multi-echo data (p=0.403; Suppl. Fig. 7e,f), although this nonrandomized comparison was not designed to establish comparative efficacy. At the end of treatment, change in HAM-$D_{17}$ did not differ between participants targeted using single-echo (N = 43) and multi-echo (N = 28) data (t = -0.842, p = 0.403). Change in MADRS also did not differ between targeting methods (t = -0.512, p = 0.610).

*Self-reported clinical outcomes*

Longitudinal change in self-reported symptom scales is reported in Suppl. Table 2, with the overall effect of treatment across post-treatment timepoints in Suppl. Table 3, and shown in Suppl. Figs. 9 and 10. Self-reported depression, suicidality, and anxiety significantly improved, whereas irritability improved less consistently and anhedonia and alexithymia did not change significantly. Contrasts testing overall post-treatment change were significant for BDI (estimate -6.66 [SE 0.92], d.f. = 66.85, t = -7.20, $p_{adj} < 0.001$), PHQ-9 (estimate -4.63 [SE 0.66], d.f. = 67.27, t = -7.01, $p_{adj} < 0.001$), QIDS-SR-16 (estimate -6.00 [SE 0.69], d.f. = 67.93, t = -8.65, $p_{adj} < 0.001$), BSS (estimate -2.60 [SE 0.52], d.f. = 65.50, t = -4.98, $p_{adj} < 0.001$), GAD-7 (estimate -2.90 [SE 0.52], d.f. = 68.00, t = -5.43, $p_{adj} < 0.001$), and BITe (estimate -1.44 [SE 0.42], d.f. = 67.75, t = -3.38, $p_{adj} = 0.012$) (Suppl. Table 3). Conversely, contrast estimates were not significant for DARS (estimate 3.83 [SE 2.04], d.f. = 67.34, t = 1.88, $p_{adj} = 0.651$) or TAS-20 (estimate -1.85 [SE 0.82], d.f. = 67.62, t = -2.26, $p_{adj} = 0.270$). In the most densely sampled measure, the BDI, improvement was significant on the first day of treatment (mean ΔBDI = -1.65 points [SE 0.56], t = -2.94, d.f. = 68, $p_{adj} = 0.044$) and continued thereafter (Suppl. Fig. 9a, Suppl. Table 2). Change in self-reported outcomes did not differ between TRD and UTRD groups on any scale at any timepoint.

*Tolerability and adverse events*

Adverse events are reported in full in Suppl. Table 4. No severe or serious adverse events occurred. The most frequent were stimulation-associated scalp discomfort (155 events affecting 47/71 patients), headache (48 events, 25/71), fatigue (33 events, 18/71), and stimulation-associated facial twitching (29 events, 14/71 patients). In an exploratory analysis, self-reported pain and discomfort decreased over treatment from 3.59/10 on the first day of treatment, and 2.87/10 on the last day (contrast, day 5 - day 1: -0.745 [SE 0.174], d.f = 69.00, t = -4.28, p < 0.001; Suppl. Fig. 11b).

During interleaved iTBS-fMRI, moderate stimulation-associated discomfort was reported by 16.2% of patients (Suppl. Table 4). In the subset with pain and discomfort ratings, scores were greater during active than sham iTBS-fMRI (t = 5.96, d.f. = 37, p < 0.001), although only 59.6%

of participants guessed the stimulation condition correctly (34/57, $p = 0.185$). Pain and discomfort during interleaved iTBS-fMRI was also greater than the average reported on the first day of open-label iTBS ($t = 2.40$, d.f. = 37, $p = 0.022$).

*Technical performance of the interleaved iTBS-fMRI setup*

Temporal signal-to-noise ratio (tSNR) during interleaved iTBS-fMRI is shown by condition and coil configuration in Suppl. Figs. 1 and 2, and the predicted reduction in E-field magnitude during sham stimulation in Suppl. Fig. 5. Across the bilateral sgACC and dlPFC E-field, as well as the whole-brain, tSNR was significantly higher during sham than active stimulation and with the later, higher-channel coil configuration (Suppl. Fig. 2). Large motion spikes were common during the first train of iTBS and occasionally in individual subjects throughout the remainder (Suppl. Fig. 13). Mean framewise displacement during iTBS-fMRI was 0.742 +/- 0.094 mm (Suppl. Fig. 13).

*Additional iTBS-fMRI analyses*

The iTBS-fMRI analysis cohort comprised 57 patients (34 TRD, 23 UTRD; Suppl. Fig. 6), with baseline characteristics reported in Suppl. Table 5 and clinician-rated outcomes in Suppl. Table 6 and Suppl. Fig. 12.

In the sham condition, seedmap responses were correlated with subsequent HAM-$D_{17}$ change at week 12 ($r = -0.324$, $p = 0.019$), but not at day 5 ($r = -0.231$, $p = 0.084$) or week 4 ($r = -0.219$, $p = 0.108$). Active-sham seedmap responses were not correlated with HAM-$D_{17}$ change at day 5 ($r = -0.078$, $p = 0.565$), week 4 ($r = -0.124$, $p = 0.367$), or week 12 ($r = -0.085$, $p = 0.550$) (Suppl. Fig. 14). No significant relationships were observed between change in HAM-$D_{17}$ and responses in the left dlPFC E-field or bilateral sgACC in the active or sham conditions as well as the active-sham contrast (Suppl. Fig. 15).

As observed in models predicting outcomes on the HAM-$D_{17}$, adding seedmap responses from the active condition to models containing age, sex, baseline MADRS severity, and MSM score improved within-sample model fit for MADRS response and remission at each follow-up timepoint (Suppl. Fig. 16). At day 5, $\Delta Dev = -9.079$ ($p = 0.003$) for response and $\Delta Dev = -10.444$ ($p = 0.001$) for remission. At week 4, $\Delta Dev = -7.613$ ($p = 0.006$) for response and $\Delta Dev = -5.329$ ($p = 0.021$) for remission. At week 12, $\Delta Dev = -4.220$ ($p = 0.040$) for response and $\Delta Dev = -6.278$ ($p = 0.012$) for remission.

Seedmap responses in the sham condition showed weaker and less consistent prognostic value. Adding sham responses improved HAM-$D_{17}$ remission model fit at day 5 ($\Delta Dev = -6.546$, $p = 0.011$) and week 12 ($\Delta Dev = -7.343$, $p = 0.007$), and MADRS remission at week 12 ($\Delta Dev = -4.154$, $p = 0.042$). Other sham-condition nested-model comparisons were not significant (Suppl. Fig. 17). Responses in the bilateral sgACC and left dlPFC E-field did not significantly improve the nested models for HAM-$D_{17}$ or MADRS response or remission at any follow-up timepoint (Suppl. Figs. 18 and 19).
